# The impact of opioid-free analgesia on pain severity and patient satisfaction after discharge from surgery: a multi-specialty, prospective cohort study in 25 countries

**DOI:** 10.1101/2023.10.01.23296409

**Authors:** William Xu, TASMAN Collaborative

## Abstract

**Background:** Balancing opioid stewardship and the need for adequate analgesia following discharge after surgery is challenging. Concern about inadequate analgesia after discharge contributes to excessive opioid prescribing, but the benefits of opioid prescription following discharge remains unclear. This study aimed to compare the outcomes for patients discharged with opioid versus opioid-free analgesia after common surgical procedures.

**Methods:** This international, multicentre, prospective cohort study collected data from patients undergoing common acute and elective general surgical, urological, gynaecological, and orthopaedic procedures. The primary outcomes were patient-reported time in severe pain during the first week following discharge, and patient-reported satisfaction with pain relief 7 days following discharge. Secondary outcomes included patient-reported quality of life, representations to healthcare for inadequately treated pain, and representations for side effects of pain medication. Data were collected by in-hospital chart review and patient telephone interview one week after discharge. Mixed-effects multivariate models, adjusted for patient comorbidity, operative characteristics, postoperative factors, country, and centre, with and without propensity score matching, were used to analyse outcomes.

**Findings:** The study recruited 4,273 patients from 144 centres in 25 countries. Overall, 1311 patients (30.7%) were prescribed opioid analgesia at discharge. Patients reported being in severe pain for 10% (IQR 1 to 30%) of the first week after discharge and rated satisfaction with analgesia as 9/10 (IQR 8 to 10). On negative binomial regression, opioid analgesia on discharge was independently associated with increased pain severity (risk ratio=1.52, 95% CI 1.31 to 1.76, p<0.001) but not with analgesia satisfaction (beta coefficient=0.92, 95% CI −1.52 to 3.36, p=0.468) when compared to opioid-free analgesia. Opioid analgesia on discharge was associated with an increased risk of representation to healthcare providers for medication side effects (OR 2.38 95%CI 1.36 to 4.17, p=0.004). While opioid prescribing varied dramatically between high income and low and middle income countries, patient reported outcomes did not.

**Interpretation:** Opioid analgesia prescription on discharge is not associated with decreased pain severity or satisfaction with analgesia after surgical discharge, but is associated with higher risk of representation for medication side effects. For many operations, opioid-free analgesia at surgical discharge should be routinely adopted without concern for uncontrolled pain or reduced patient satisfaction.

**What this study adds:** *Evidence before this study:* We conducted a literature search between November 2019 and February 2021 for studies on the relationship between opioid prescription and patient reported satisfaction or pain after discharge from surgery. We searched MEDLINE, Google Scholar and ClinicalTrials.gov using the search terms “opioid”, “surgery”, “discharge” without any language restrictions. Several single centre and retrospective surgical series examined opioid prescription practices after surgery demonstrating overprescription. Global studies examining variations in opioid prescribing between countries are less common but demonstrate significant global variation in prescription practices. One recent systematic review and meta-analysis examined randomised controlled trials comparing opioid and opioid-free analgesia in the post-surgical-discharge, and showed no difference between the two groups, but was limited to elective minor and moderate surgical procedures.

*Added value of this study:* This large study provides patient-reported data on pain and patient satisfaction after discharge from surgery. This adds to the previous knowledge by including both minor and major operations in an acute and elective context, and multiple specialties. After adjustment, opioid analgesia on discharge was not associated with decreased time in severe pain or increased patient satisfaction, but was associated with an increased risk of re-presentation for medication side effects. We also demonstrate marked geographical variation in opioid prescribing practices with higher amounts prescribed in high income countries compared to low and middle income countries, without a similar variation in patient reported pain or satisfaction.

*Implications of all the available evidence:* Opioids are often prescribed at the time of discharge from hospital following surgery, but the benefit of post-discharge opioids has been called into question. We found that that opioids do not reduce severity of pain during the first week after discharge and do not increase patient satisfaction. These data suggest that opioid-free analgesia at surgical discharge is feasible without the risk of increased pain or decreased satisfaction, and that opioids should be prescribed more selectively. Variation in opioid prescribing between countries is not associated with variations in pain or satisfaction, and suggests that a more uniform approach to analgesia prescribing is warranted.

## Introduction

Postoperative pain is common, complex, and often severe, affecting up to 80% of patients.^1^ It is frequently managed using opioid analgesia, which although potent, incurs the risk of significant adverse events. There are ongoing concerns regarding the inappropriate, excessive, and unsafe prescription of opioid analgesia after surgery.^2, 3^ Such practices are associated with worse patient outcomes, greater healthcare demands, and the ‘opioid epidemic’.^4, 5^

Surgeons are high prescribers of opioids, accounting for approximately 10% of all prescriptions.^6^ The overprescription of opioids in the postoperative period likely stems from healthcare providers’ desire to reduce patient discomfort, and concerns regarding poor patient satisfaction.^7, 8^ Unused postoperative opioids represent an important contributor to opioid diversion in the community.^9^ Despite the perception that opioids aid pain management in this setting, multiple studies have indicated that opioid-sparing protocols following surgery are not associated patient satisfaction, provided that pain is adequately controlled using non-opioid analgesia, and patient expectations are appropriately managed.^7,10^

A recent meta-analysis by Fiore Jr *et al.* reported that opioid prescribing at discharge from elective procedures did not improve pain control, but was associated with increased harm.^11^ This review was limited to minor and moderate procedures such as dental procedures or cholecystectomies, but demonstrated that the default stance to prescribe opioids at surgical discharge may be a practice steeped in culture rather than evidence. Given the variation in prescribing practices between countries, centres, and individual clinicians,^12^ international, multi-specialty data are required to understand the relationship between opioid prescription and patient-reported pain and satisfaction outcomes, particularly following emergency and major procedures.

This study aimed to describe patient-reported outcomes of patient pain, satisfaction, and quality of life after common surgical procedures, and investigate the effect of opioid prescription on patient-reported post-discharge outcomes and risk of representation for inadequate analgesia or adverse effects.

## Methods

### Study design

The Opioid PrEscRiptions and Usage After Surgery (OPERAS) study was an international multi-centre collaborative study developed by the Trials and Audit in Surgery by Medical Students in Australia and New Zealand (TASMAN) Collaborative, an Australasian student- and trainee-led collaborative network. The study protocol (**Appendix S1**) has been published elsewhere.^13^ The collaborative research model has been used by other studies internationally and has been described previously.^14, 15^ Study requirements and approvals were achieved according to country-specific regulations before participant recruitment began. This analysis was conducted in line with the STROBE reporting guidelines for observational studies.^16^

### Ethical approval

Ethical approval was obtained at each participating site in line with local protocols and verified by the central steering committee. The protocol was approved by the Hunter New England Human Research Ethics Committee (2021/ETH11508) in Australia as the lead site.

### Eligibility criteria

Adult patients (aged 18 years or above) who underwent an eligible general surgical (cholecystectomy, appendicectomy, inguinal hernia repair, colon resection, fundoplication, or sleeve gastrectomy), orthopaedic (total or reverse shoulder arthroplasty, rotator cuff or labral repair, anterior cruciate ligament repair, or hip or knee arthroplasty), gynaecological (hysterectomy, oophorectomy, or salpingectomy and oophorectomy), or urological operation (prostatectomy, cystectomy or nephrectomy) during the data collection periods were approached for inclusion.^13^ Elective and emergency surgeries were included. Patients who fulfilled any of the following criteria were excluded: 1) currently on medication-assisted treatment of opioid dependence; 2) discharged to another healthcare setting (e.g. rehabilitation service); 3) multivisceral resection; 4) returned to the operating room during their index admission. Eligible patients were identified through inspection of surgical operating lists. All participants provided informed consent prior to inclusion and each centre obtained ethical approval prior to data collection.

### Data collection

Data were prospectively collected during six separate two-week periods between April and September 2022, from inpatient hospital records and by a follow-up phone call at 7 days post-discharge. The follow-up interview was conducted using a pre-specified protocol and script (**Appendix S2**) to ensure standardisation.

Data were collected on patient demographics (age, Sex, smoking status, BMI, Society of Anesthesiologists (ASA) physical status classification), comorbidities, diagnosis and procedure specific details (indication, surgical approach, and urgency), opioid use in the 24 hours prior to hospital discharge, post-operative complications, preoperative regular opioid and non-opioid analgesic use. Regular analgesic use was defined as a minimum of once-weekly use in the 3 months immediately prior to surgery. Minimally invasive surgery was defined as arthroscopic, laparoscopic or robotic surgery. Open surgery was defined as planned open, and laparoscopic/robotic converted to open procedures. Data were also collected on opioid and non-opioid medication prescriptions at discharge.

To enable comparison between opioids of different potencies, all data on opioid doses were converted to oral morphine equivalents (OMEs). Further details on how OMEs were calculated are provided in **Appendix S3**.

### Outcomes

The primary outcome was the amount of time spent in severe pain in the first 7 days following discharge measured on a 0 to 100% numerical analogue scale. Secondary outcomes included patient-reported satisfaction with the quality of analgesia received on a scale of 0 to 100, patient-reported quality of life measured by the EQ-5D-5L tool 7 days after discharge, number of presentations to healthcare professionals for inadequate analgesia, and number of presentations to healthcare for side effects of analgesia including nausea, vomiting, drowsiness, itching, dizziness or constipation. Outcome measures related to the analgesic prescription and consumption are reported elsewhere.<u>(OPERAS Steering Committee 2023)</u>

The EQ-5D-5L tool is used to measure patient quality of life in five domains including mobility, self-care, usual activities, pain/discomfort and anxiety/depression. The tool also includes an EQ-VAS score, which rates a patient’s self-reported health from 0 (worst possible) to 100 (best possible).^17^ Re-presentations to healthcare were defined as any visit to primary care, emergency departments, the surgeon’s office, or readmission to hospital for inadequately treated pain or side effects of analgesic medication between discharge and 7-days post-discharge.

### Statistical analysis

Analyses were completed in R 4.0.3 for statistical computing (Vienna, Austria. R Core Team). Descriptive statistics were used to compare demographic and in-hospital differences between patients discharged on opioid and opioid-free analgesia using χ2 tests for categorical variables and Kruskal-Wallis tests for continuous variables.

Propensity score matching was used to minimise the selection bias of participants prescribed opioid and opioid-free analgesia on discharge using the *MatchIt* package.^18^ The propensity score was defined as the probability that a patient would receive opioid analgesia adjusted for age, sex, ASA, BMI, presence of chronic kidney disease and liver disease, smoking status, preoperative opioid and non-opioid analgesia use, surgical procedure, duration, surgical indication, postoperative complications, length of stay, concomitant discharge paracetamol and NSAID prescription, and OMEs consumed in the 24 hours prior to discharge. Full matching was used to allow multiple patients from each group to be matched together (if appropriate) and weighted for balancing, avoiding inappropriate discarding of data that can occur with nearest-neighbour matching.^19^ Balance between groups was assessed before and after using the standardised mean difference, with an absolute value of less than 0.1 as an indication of a well-balanced variable.

Multiple imputation by chained equations was used to impute values for patients with missing data using the *mice* package.^20^ Visual inspection of variables with missing data stratified by presence of opioid analgesia at discharge was completed to ensure variables were missing at random. Ten imputed datasets were created with propensity score matching subsequently performed on each using the *MatchThem* package.^21^ The pooled results of models are presented.

Mixed-effects models, using centre and country as random effects, were fitted for primary and secondary outcomes. Logistic regression models were built for binary outcomes, and negative binomial regression or generalised linear regression models were built for continuous outcomes. Covariate selection was guided by clinical plausibility and previous literature,^22^ and relevant preoperative, intraoperative, and postoperative variables were included as fixed effects. Residual, Q-Q plots, and variance inflation factors were interrogated to assess model assumptions.

### Subgroup analysis

Procedures were stratified into abdominal procedures (including general, gynaecological, and urological operations) and orthopaedic procedures. Sensitivity analyses were performed on these subgroups. Prescribing practices and patient reported outcomes were compared between different regions of the world, and between high income countries (HIC) and low and middle income countries (LMIC) as defined by Organisation for Economic Co-operation and Development (OECD).^23^

## Results

Over the study period, data from 4273 patients from 114 hospitals in 25 countries were collected (**Figure 1**). The majority of patients included were from Australia (n = 813), Egypt (n = 594), Aotearoa New Zealand (n = 560), Libya (n = 372), and Turkey (n = 296); the full contributions from each country are listed in **Table S1**. Patient demographics and comorbidities stratified by opioids versus no opioids on discharge are presented in **Table 1** (median age 50 years, 53.1% female). The majority of included patients underwent general surgical procedures (71.5%, n = 3056), followed by orthopaedic procedures (13.8%, n = 591), with a minority of gynaecological (9.2%, n = 393) and urological procedures (5.5%, n = 233; **Table 1**).

**Figure 1.**
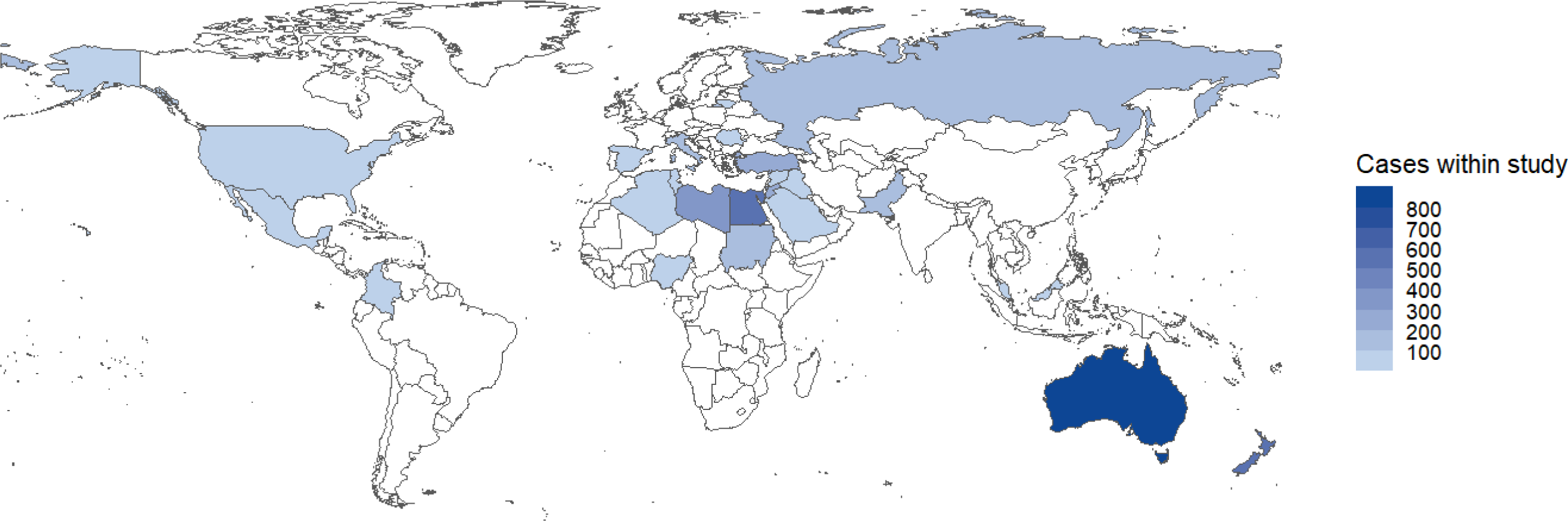
Cases contributed to the OPERAS study by country. Full details in Table S1

**Table 1.**

| Variable |  | No opioids | Opioids | Total | p |
| --- | --- | --- | --- | --- | --- |
| Total N (%) |  | 2962 (69.3) | 1311 (30.7) | 4273 |  |
| Age | Median (IQR) | 48.0 (33.0 to 64.0) | 52.0 (37.0 to 66.0) | 50.0 (34.0 to 64.0) | <0.001 |
| Sex | Female | 1579 (53.3) | 692 (52.8) | 2271 (53.1) | 0.033 |
|  | Male | 1383 (46.7) | 616 (47.0) | 1999 (46.8) |  |
|  | Other |  | 3 (0.2) | 3 (0.1) |  |
| ASA | I | 1377 (46.5) | 386 (29.4) | 1763 (41.3) | <0.001 |
|  | II-III | 1559 (52.6) | 907 (69.2) | 2466 (57.7) |  |
|  | IV - V | 24 (0.8) | 15 (1.1) | 39 (0.9) |  |
|  | (Missing) | 2 (0.1) | 3 (0.2) | 5 (0.1) |  |
| BMI | Normal (18.5 -24.9) | 835 (28.2) | 327 (24.9) | 1162 (27.2) | <0.001 |
|  | Overweight (25-30) | 1078 (36.4) | 385 (29.4) | 1463 (34.2) |  |
|  | Obese (31-40) | 603 (20.4) | 333 (25.4) | 936 (21.9) |  |
|  | Severely obese (>40) | 111 (3.7) | 94 (7.2) | 205 (4.8) |  |
|  | Underweight (< 18.5) | 56 (1.9) | 13 (1.0) | 69 (1.6) |  |
|  | (Missing) | 279 (9.4) | 159 (12.1) | 438 (10.3) |  |
| MI or CHF | No | 2808 (94.8) | 1227 (93.6) | 4035 (94.4) | 0.13 |
|  | Yes | 154 (5.2) | 84 (6.4) | 238 (5.6) |  |
| PVD | No | 2877 (97.1) | 1276 (97.3) | 4153 (97.2) | 0.791 |
|  | Yes | 85 (2.9) | 35 (2.7) | 120 (2.8) |  |
| CVA or TIA | No | 2903 (98.0) | 1285 (98.0) | 4188 (98.0) | 1 |
|  | Yes | 59 (2.0) | 26 (2.0) | 85 (2.0) |  |
| PUD | No | 2900 (97.9) | 1285 (98.0) | 4185 (97.9) | 0.907 |
|  | Yes | 62 (2.1) | 26 (2.0) | 88 (2.1) |  |
| Diabetes | No | 2559 (86.4) | 1115 (85.0) | 3674 (86.0) | 0.263 |
|  | Yes | 403 (13.6) | 196 (15.0) | 599 (14.0) |  |
| CKD | No | 2913 (98.3) | 1268 (96.7) | 4181 (97.8) | 0.001 |
|  | Yes | 49 (1.7) | 43 (3.3) | 92 (2.2) |  |
| Liver disease | No | 2910 (98.2) | 1283 (97.9) | 4193 (98.1) | 0.47 |
|  | Yes | 52 (1.8) | 28 (2.1) | 80 (1.9) |  |
| Comorbid Cancer | No | 2749 (92.8) | 1153 (87.9) | 3902 (91.3) | <0.001 |
|  | Yes | 213 (7.2) | 158 (12.1) | 371 (8.7) |  |
| Smoking | Never smoked | 1841 (62.2) | 723 (55.1) | 2564 (60.0) | <0.001 |
|  | Current smoker | 550 (18.6) | 182 (13.9) | 732 (17.1) |  |
|  | Ex-smoker | 355 (12.0) | 307 (23.4) | 662 (15.5) |  |
|  | (Missing) | 216 (7.3) | 99 (7.6) | 315 (7.4) |  |
| Procedure | Appendicectomy | 522 (17.6) | 241 (18.4) | 763 (17.9) | <0.001 |
|  | Cholecystectomy | 880 (29.7) | 346 (26.4) | 1226 (28.7) |  |
|  | Colorectal resection | 271 (9.1) | 122 (9.3) | 393 (9.2) |  |
|  | Inguinal hernia repair | 440 (14.9) | 136 (10.4) | 576 (13.5) |  |
|  | Nissen fundoplication | 23 (0.8) | 5 (0.4) | 28 (0.7) |  |
|  | Sleeve gastrectomy | 49 (1.7) | 21 (1.6) | 70 (1.6) |  |
|  | ACL repair | 57 (1.9) | 21 (1.6) | 78 (1.8) |  |
|  | Knee arthroplasty | 110 (3.7) | 147 (11.2) | 257 (6.0) |  |
|  | Hip arthroplasty | 110 (3.7) | 91 (6.9) | 201 (4.7) |  |
|  | Rotator cuff repair | 11 (0.4) | 11 (0.8) | 22 (0.5) |  |
|  | Shoulder arthroplasty | 9 (0.3) | 11 (0.8) | 20 (0.5) |  |
|  | Shoulder labral repair | 12 (0.4) | 1 (0.1) | 13 (0.3) |  |
|  | Cystectomy | 28 (0.9) | 4 (0.3) | 32 (0.7) |  |
|  | Nephrectomy | 62 (2.1) | 38 (2.9) | 100 (2.3) |  |
|  | Prostatectomy | 66 (2.2) | 35 (2.7) | 101 (2.4) |  |
|  | Hysterectomy | 228 (7.7) | 59 (4.5) | 287 (6.7) |  |
|  | Oophorectomy and Salpingectomy | 28 (0.9) | 13 (1.0) | 41 (1.0) |  |
|  | Oophorectomy only | 21 (0.7) | 3 (0.2) | 24 (0.6) |  |
|  | Salpingectomy only | 35 (1.2) | 6 (0.5) | 41 (1.0) |  |
| Surgical approach | MIS | 1487 (50.2) | 814 (62.1) | 2301 (53.8) | <0.001 |
|  | Open | 1474 (49.8) | 496 (37.8) | 1970 (46.1) |  |
|  | (Missing) | 1 (0.0) | 1 (0.1) | 2 (0.0) |  |
| Indication | Benign | 2601 (87.8) | 1144 (87.3) | 3745 (87.6) | 0.63 |
|  | Malignancy | 360 (12.2) | 167 (12.7) | 527 (12.3) |  |
|  | (Missing) | 1 (0.0) | 0 (0.0) | 1 (0.0) |  |
| Urgency | Elective | 2098 (70.8) | 818 (62.4) | 2916 (68.2) | <0.001 |
|  | Emergency | 863 (29.1) | 493 (37.6) | 1356 (31.7) |  |
|  | (Missing) | 1 (0.0) | 0 (0.0) | 1 (0.0) |  |
| Surgical duration (min) | Median (IQR) | 80.0 (55.0 to 120.0) | 98.0 (66.0 to 135.0) | 87.0 (60.0 to 120.0) | <0.001 |
| Opioid use pre-surgery | No | 2909 (98.2) | 1180 (90.0) | 4089 (95.7) | <0.001 |
|  | Yes | 53 (1.8) | 131 (10.0) | 184 (4.3) |  |
| Non-opioid analgesia use pre-surgery | No | 2396 (80.9) | 933 (71.2) | 3329 (77.9) | <0.001 |
|  | Yes | 566 (19.1) | 378 (28.8) | 944 (22.1) |  |
| Clavien-Dindo complication | None | 2450 (82.7) | 1025 (78.2) | 3475 (81.3) | <0.001 |
|  | I-II | 487 (16.4) | 265 (20.2) | 752 (17.6) |  |
|  | III-IV | 21 (0.7) | 20 (1.5) | 41 (1.0) |  |
|  | (Missing) | 4 (0.1) | 1 (0.1) | 5 (0.1) |  |
| Length of stay (days) | Median (IQR) | 2.0 (1.0 to 3.0) | 2.0 (1.0 to 3.0) | 2.0 (1.0 to 3.0) | 0.698 |
| OMEs consumed 24h before discharge | Median (IQR) | 0.0 (0.0 to 30.0) | 35.0 (10.0 to 67.8) | 9.0 (0.0 to 40.0) | <0.001 |

A total of 30.7% of patients (n = 1311) were prescribed opioid analgesia at discharge. For patients prescribed any opioid on discharge, the median quantity prescribed was 100 OMEs (IQR 60 to 200). However, at 7-days follow-up the median quantity consumed by patients was only 40 OME (IQR 7.5 - 100; p<0.001). Complete data on the proportion of prescribed opioids that were consumed by 7 days are described elsewhere.<u>(OPERAS Steering Committee 2023)</u> Of note, 15.0% (n = 197) of patients who received a prescription for opioids at discharge did not consume any opioid analgesia in the 24 hours prior to discharge. A total of 60.8% (n = 2596) patients recalled receiving education about pain management prior to discharge.

Propensity score matching produced well matched cohorts, demonstrated by the plot of standardised mean differences of included variables in **Figure S1** and **Table S2.**

### Post-discharge patient reported outcomes

The reported time spent in severe pain after discharge, stratified by procedure, is displayed in **Figure 2A**. On univariate analysis of the overall cohort, there was a statistically significant difference in the time spent in severe pain after discharge between those discharged with and without opioids (median: 20% prescribed opioids vs 10% no opioids, p<0.001; **Table 2**). After propensity score matching and adjustment for confounding factors on mixed-effects negative binomial regression, patients prescribed opioids spent more time in severe pain after discharge (RR 1.52, 95%CI 1.31 to 1.76, p < 0.001, **Table 3, Table S3**).

**Figure 2.**
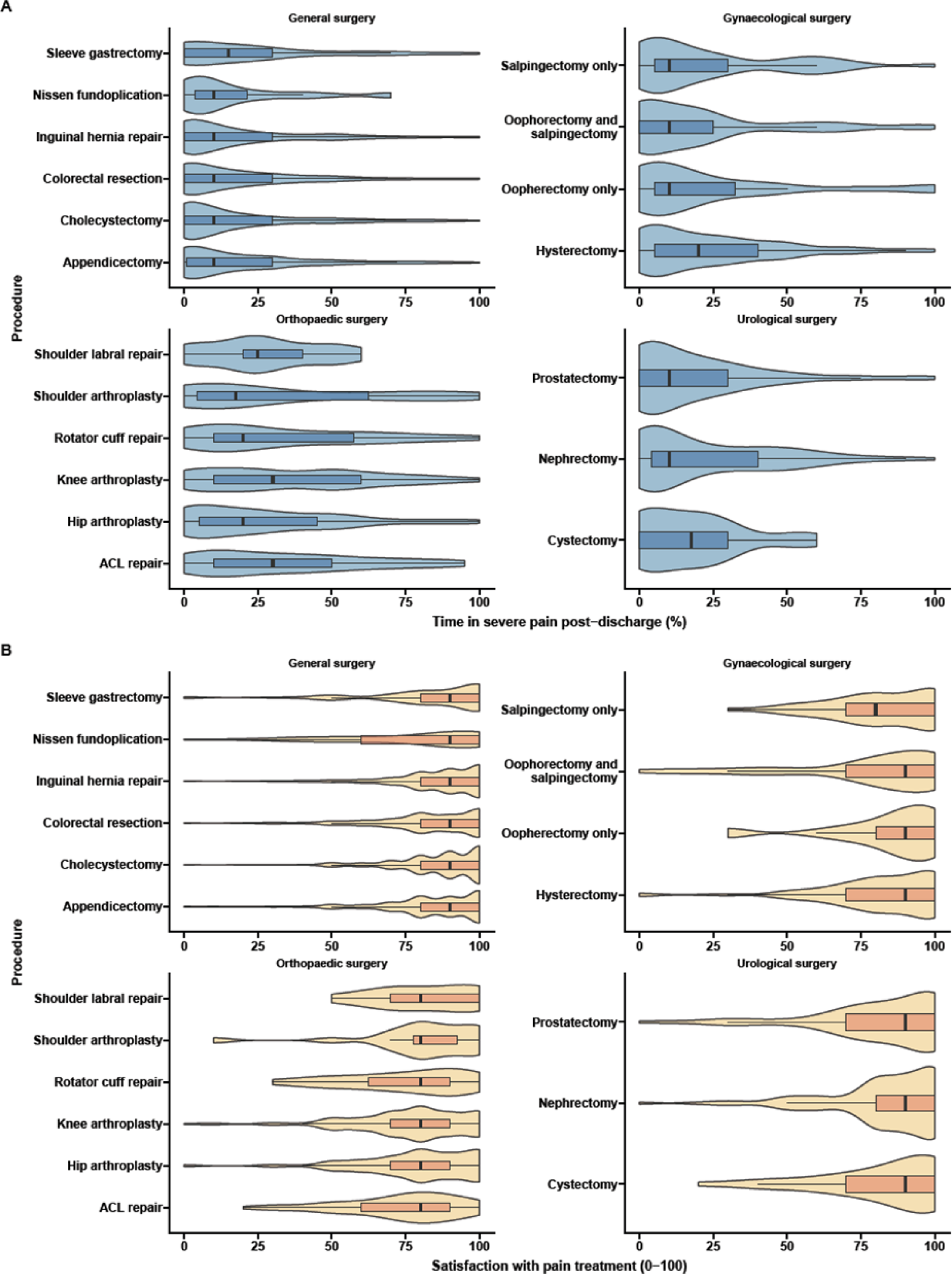
Outcomes of pain severity post discharge and satisfaction with pain treatment stratified by procedure. ACL: anterior cruciate ligament

**Table 2.**

| Variable |  | No opioids | Opioids | Total | p |
| --- | --- | --- | --- | --- | --- |
| **Total N (%)** |  | **2962 (69.3)** | **1311 (30.7)** | **4273** |  |
| Percent time in severe pain in week post-discharge (%) | Median (IQR) | 10.0 (0.0 to 30.0) | 20.0 (5.0 to 40.0) | 10.0 (1.0 to 30.0) | <0.001 |
| Satisfaction with pain treatment (0-100) | Median (IQR) | 90 (80 to 100) | 90 (80 to 100) | 90 (80 to 100) | 0.157 |
| Satisfaction amount of analgesia provided | Too little | 622 (21.0) | 229 (17.5) | 851 (19.9) | <0.001 |
|  | Just right | 2226 (75.2) | 887 (67.7) | 3113 (72.9) |  |
|  | Too much | 110 (3.7) | 194 (14.8) | 304 (7.1) |  |
|  | (Missing) | 4 (0.1) | 1 (0.1) | 5 (0.1) |  |
| EQ-5D-5L composite score (0-100) | Median (IQR) | 87.9 (76.1 to 95.0) | 80.9 (69.1 to 89.2) | 85.9 (73.8 to 93.7) | <0.001 |
| EQ-VAS score (0-100) | Median (IQR) | 82.0 (70.0 to 90.0) | 75.0 (60.0 to 85.0) | 80.0 (70.0 to 90.0) | <0.001 |
| Post-discharge medical presentation for pain | No | 2701 (91.2) | 1121 (85.5) | 3822 (89.4) | <0.001 |
|  | Yes | 261 (8.8) | 190 (14.5) | 451 (10.6) |  |
| Post-discharge medical presentation for medication SEs | No | 2851 (96.3) | 1220 (93.1) | 4071 (95.3) | <0.001 |
|  | Yes | 111 (3.7) | 91 (6.9) | 202 (4.7) |  |

**Table 3.**
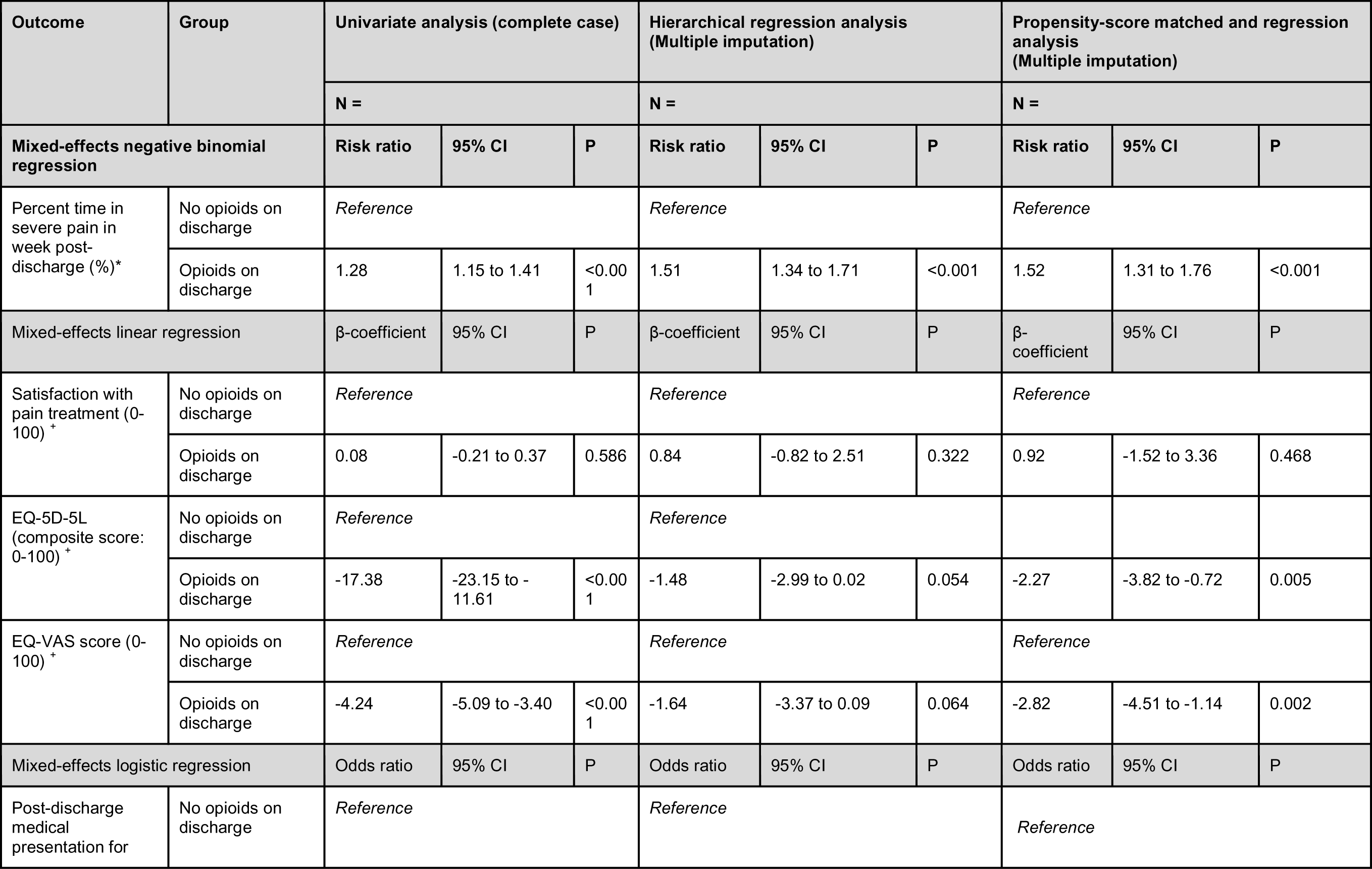
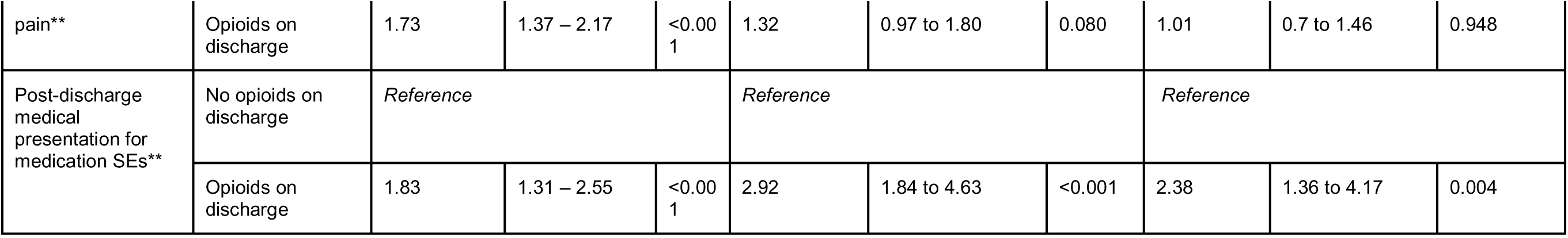

Despite the differences in pain severity for patients discharged with and without opioids, there was no difference in patient-reported satisfaction with pain treatment on univariate analysis (median satisfaction rating: 90/100 opioids vs 90/100 no opioids, p = 0.157, **Table 2**). Further, no significant association was found between opioid analgesia and patient satisfaction score after propensity score matching and adjustment for confounding factors on mixed-effects linear regression (β = 0.92, 95%CI −1.52 to 3.36, p = 0.468, **Table 3, Table S4**).

There was no dose-dependent relationship between the quantity of opioids prescribed on discharge and pain severity or patient reported satisfaction on both unadjusted (**Figure S2**) and adjusted analyses (**Figure 3**).

**Figure 3.**
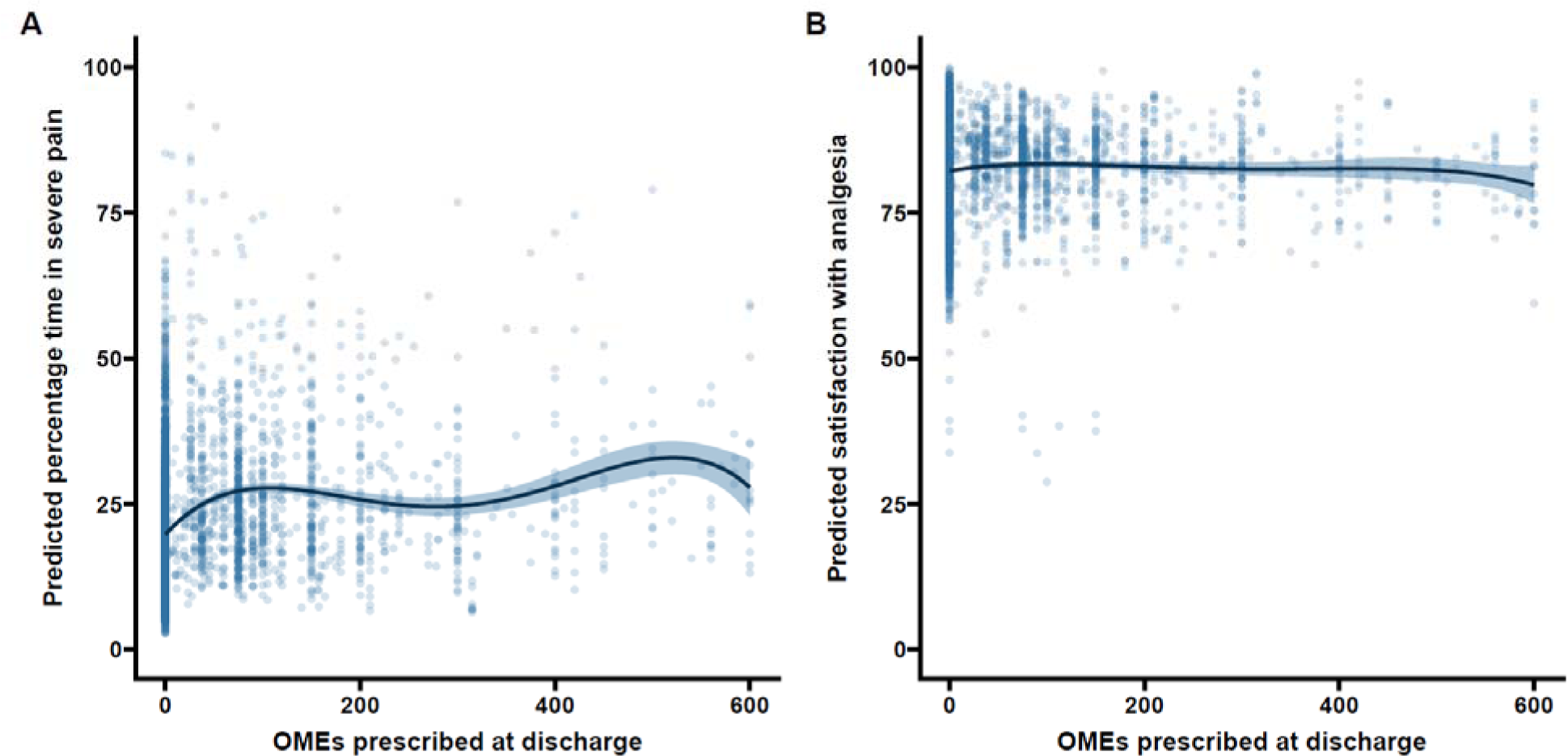
Relationship between quantity of OMEs prescribed at discharge and A) modelled time in severe pain and B) modelled patient satisfaction with pain treatment adjusted for patient demographics, comorbidity, operative type, duration and indication, preoperative opioid and non-opioid analgesia, postoperative complications, and OME requirements 24h prior discharge.

### Quality of life

Before risk adjustment, patients prescribed opioids reported lower quality of life at 7 days post-discharge compared to those not prescribed opiates when measured by a composite EQ-5D-5L score (80.9 vs 87.9, p<0.001) or EQ-VAS score (75.0 vs 82.0, p<0.001). After adjustment on mixed-effects linear regression and propensity-score matching, opioid prescription on discharge was associated with lower quality of life measured by both EQ-5D-5L score (β = −2.27, p=0.005, **Table 3, Table S5**) and EQ-VAS score (β = −2.82, p=0.002, **Table 3, Table S6**).

### Re-presentation to healthcare

A total of 451 (10.6%) of patients sought additional pain relief after hospital discharge, while 202 patients (4.7%) presented to healthcare due to medication side effects like nausea, constipation, or pruritus. On adjusted analysis after propensity score matching, opioid prescriptions at discharge did not increase the likelihood of patients seeking additional healthcare for pain relief (OR 1.01, 95% CI 0.70 to 1.46, p = 0.948, Table 3, Table S7), but increased the risk of presentation to healthcare due to side effects of medication (OR 2.38, 95% CI 1.36 to 4.17, p= 0.004, Table 3, Table S8).

### Excess and insufficient analgesia prescription

Overall, 14.8% of patients prescribed opioid analgesia felt they were prescribed too much pain relief medication compared to 3.7% of those prescribed only non-opioid analgesia. Conversely, 17.5% of patients prescribed opioids felt they were prescribed too little pain relief medication compared with 21.0% of those prescribed no opioids (p < 0.001, **Table 2**).

Factors associated with patients reporting receiving too little pain relief on multivariate analysis included female sex (OR 1.25 95% CI 1.03 to 1.51, p<0.001), preoperative regular use of opioid analgesia (OR 1.94 95% CI 1.32 to 2.82, p<0.001), ASA IV-V (OR 2.66 95% CI 1.27 to 5.54 compared to ASA I) and postoperative complications (Clavien Dindo Grade I-II OR 1.41 95% CI 1.13 to 1.76, p<0.003 compared to no complications). Patients who had undergone orthopaedic procedures of ACL repair (OR 2.83 95% CI 1.53 to 5.24, p<0.001), knee arthroplasty (OR 2.37 95% CI 1.43 to 3.93, p<0.001) or shoulder arthroplasty (OR 4.39 95% CI 1.50 to 12.91, p<0.001) or cystectomy (OR 3.06 95% CI 1.22 to 7.68) were also more likely to report receiving too little pain relief compared to patients who had undergone appendectomy. Conversely, patients prescribed opioids (OR 0.59 95% CI 0.46 to 0.75, p<0.001) or paracetamol on discharge (OR 0.65 95% CI 0.52 to 0.81, p<0.001) were less likely to think they received too little pain medication (**Table S9**).

### Subgroup analysis: impact of surgical procedure

Patients who underwent orthopaedic procedures reported severe pain more frequently in the 7 days following discharge when compared to those who underwent abdominal procedures (**Figure 2A**, orthopaedic 30% IQR: 10 to 50 vs abdominal 10% IQR: 0 to 30, p <0.001), and reported a lower level of satisfaction with pain relief post discharge (**Figure 2B**, orthopaedic 80/100 IQR: 70 to 90 vs abdominal 90/100 IQR: 80 to 100, p<0.001). Orthopaedic patients reported lower quality of life scores post-discharge (orthopaedic EQ-VAS 70/100 IQR: 60 to 80 vs abdominal 80/100 IQR 70 to 90, p<0.001), were more likely to seek further analgesia (18.3% vs 9.3%, p<0.001), and were more likely to represent to healthcare for side effects of analgesia (9.1% vs 4.0%, p<0.001).

Within the orthopaedic cohort, prescription of opioids on discharge did not have a significant impact on outcomes: pain severity post discharge (RR 1.17 95% CI 0.84 to 1.53, p = 0.410), satisfaction with pain treatment (β = 3.01, 95% CI −1.69 to 7.72, p < 0.001), EQ-VAS quality of life (β = −3.39, 95%CI −7.3 to 0.52, p = 0.092), risk of presentation for pain (OR 0.93 95% CI 0.42 to 2.08, p = 0.868), or risk of presentation for side effects of medication (OR 1.71 95% CI 0.46 to 6.32, p = 0.425).

In the abdominal surgery subgroup (general, gynaecological and urological procedures), discharge opioids were associated with increasing pain severity post discharge (RR 1.59 95% CI 1.35 to 1.87, p<0.001), worse EQ-VAS QoL (β = −1.87, 95% CI −3.82 to 0.08, p <0.001), and increased risk of presentation for side effects of medication (OR 2.74 95% CI 1.46 - 5.16, p = 0.003). However, discharge prescription of opioid analgesia were not associated with satisfaction with pain treatment (β = 1.69, 95% CI −0.46 - 3.83, p = 0.13), risk of presentation for pain (OR 1.29 95% CI 0.88 - 1.89, p = 0.203).

### Geographical variation

Patients from HIC as defined by OECD (n = 1923) were prescribed more opioids on average than patients from LMIC (n = 2350) (37.5 OME [IQR: 0 to 112.5] in HIC vs 0 OME [IQR 0 to 0] LMIC; p<0.001). Patients from HIC were at 9 times higher odds of receiving opioids on surgical discharge compared with patients from LMIC after adjusting for case mix, patient comorbidity, postoperative complications, analgesic needs after propensity score matching (adjusted OR 9.1 95% 7.7 to 11.1, **Table S10**). Similar results were found without propensity score matching (adjusted OR 10.0 95% 8.3 to 12.5).

While there was a statistically significant difference, there were no clinically significant differences in time spent in severe pain (10% in first week [IQR 0% to 30%] HIC vs 10% [3% to 30%] LMIC; p<0.001) or patient satisfaction between HIC and LMIC countries (90/100 [IQR 80 to 100] HIC vs 85/100 [IQR 70 to 100] LMIC; p<0.001).

Geographical differences in patients and outcomes between Asia Pacific, North America, Central and Latin America, Middle East and North Africa, Europe and Central Asia, Sub-Saharan Africa, and South Asia are presented in full in **Table S11** and **S12.**

## Discussion

This multinational prospective cohort study demonstrated that prescription of opioids at hospital discharge after common surgical procedures was not associated with lower pain severity or treatment satisfaction when compared to opioid-free analgesia. This effect also persisted when analysing the relationship between quantity of opioid prescribed and patient-reported outcomes. Opioid prescription after common general, urological, gynaecological and surgical procedures was associated with increased presentations to healthcare for management of pain medication side effects, without an associated reduction in presentations for further pain management. This study expands on previous work, which was largely limited to minor elective day-case procedures,^11, 24–26^ by including acute operations, major visceral resections, and major orthopaedic procedures. This study provides prospective, international data to inform discharge analgesia prescription after common surgical procedures and highlights that opioid-free analgesia at discharge can be the default rather than the exception.

It is accepted that opioid prescribing must be individualised, although reliably determining patient-needs remains challenging. It is becoming increasingly apparent across a range of surgical procedures that most patients do not benefit from opioid pain relief at discharge, with only a small targeted set of patients requiring opioid prescriptions on discharge.^11, 24, 27^ Multiple studies have attempted to identify preoperative variables associated with increased postoperative pain in order to identify a subset of patients for whom opioid prescriptions may be useful.^22^ In this study, female sex, opioid use preoperatively and on discharge, lower limb orthopaedic surgery, elective procedures, and mild postoperative complications (Clavien Dindo grades I-II) were associated with increased time spent in severe pain, which is largely consistent with findings reported in other reviews.^22, 28^ As noted by previous studies, patients using opioids preoperatively may be particularly vulnerable to uncontrolled pain and have a requirement for ongoing postoperative opioids.^29^ Our study reinforces previous findings that opioid prescription on discharge is not independently associated with reduced pain severity, and indicates that clinicians should not prescribe opioids as a panacea for postoperative pain.

Clinician concern surrounding patient dissatisfaction post-discharge and healthcare re-utilisation due to uncontrolled pain is a major driver of opioid overprescription.^7, 30^ In our cohort, both opioid prescription on discharge and quantity of opioid medication prescribed on discharge did not improve patient satisfaction at 7-days post-discharge. Furthermore, this study showed no increase in healthcare service utilisation for inadequately treated pain in the opioid-free analgesia cohort. On the contrary, healthcare utilisation for patients experiencing side effects of medication was increased for those with opioid analgesia compared to those receiving opioid-free analgesia, a finding which replicates previous studies.^11^ Similar studies conducted in laparoscopic cholecystectomy, appendectomy and minor hernia repair,^7, 10^ breast procedures,^25, 26^ more complex abdominal and urological procedures,^11, 27^ gynaecological procedures including laparotomies,^31^ as well as orthopaedic sports operations,^32^ have shown decreasing opioid prescriptions or opioid-free analgesia post discharge does not decrease patient satisfaction scores postoperatively. National surveys conducted within the United States have reported that the majority of patients preferred non-opioid over opioid analgesia if the option was available postoperatively;^1, 33^ it follows opioid-free analgesia may better align with patient wishes. Concerns regarding patient satisfaction or re-presentation to healthcare providers are therefore not valid reasons for opioid prescription on surgical discharge.

Marked global variation in opioid prescribing was demonstrated in this cohort. Similar geographical differences have been demonstrated in other studies, with the United States and Canada typically seeing significantly higher quantities of opioids being prescribed for the same procedures when compared with other countries including Sweden, China, Lebanon, Brazil, Mexico and the Netherlands.^34, 35^ This study extends the findings of previous studies and demonstrates that at 7 days post discharge follow-up, there is no clinically meaningful difference in reported pain levels when the patient cohort is stratified by geography. Current geographic variations in opioid prescribing likely reflect entrenched medico-cultural practices rather than evidence based pain management.

Patient outcomes differ across different surgical procedures, with patients who undergo orthopaedic procedures having higher postoperative pain and lower satisfaction in comparison to abdominal procedures. Given many patients undergoing arthroplasty are regular users of opioids in the pre-operative period, patients in this situation are at a higher risk of uncontrolled postoperative pain and chronic opioid use after surgery.^29^ However, regardless of procedure, a multimodal approach to pain management is required, with preoperative assessment for high-risk pain characteristics, appropriate modulation of patient expectations, ongoing pain assessment, multimodal analgesia, non-pharmacological techniques, and pain treatment planning on discharge.^36^ Preoperative counselling on pain management and opioid use may reduce patient reported pain scores, increase the likelihood of patients using non-pharmacological therapies, and increase levels of function at 6 months postoperatively.^37^ It follows that surgical teams have a duty in counselling high-risk patients on postoperative pain expectations, non-pharmacological strategies, and initiating postoperative opioid weans.

This study was able to collect prospective international patient-reported data across a range of surgical specialties with high rates of follow up. Nevertheless, there are several limitations. After extensive covariate adjustment, matching, and subgroup analysis, this study showed consistent results with other randomised data,^11^ However, observational data is not a substitute for randomised data for deducing causal relationships and results need to be interpreted with this in mind. Furthermore, although extensive physical comorbidity data was collected, this study was unable to adjust for coexisting anxiety, depression or pain catastrophizing which are other factors associated with pain severity.^22^ At follow-up, patient reported outcomes may be prone to recall bias, but this was minimised by the relatively short duration of follow-up. While a seven-day follow up may be considered a short timeframe for outcomes to develop, this was selected as clinical care standards recommend limiting the duration of usual discharge opioid prescriptions to less than 7 days.^38, 39^ Lastly, cultural differences in pain perception and reporting may confound patient recall of outcomes.^40^ We mitigated this potential risk to the results by factoring in centre and country level effects into multivariate models. Clinical judgement remains paramount to identifying patients who are at high risk of severe postoperative pain, and implementing a multimodal analgesic and pain education plan for these patients along with careful evaluation is still necessary.

In conclusion, this study suggests that opioid prescribing at surgical discharge is not associated with reduced pain intensity but is associated with increased risk of presentation to healthcare for side effects of pain medication. Opioid-free analgesia at surgical discharge should be adopted for the majority of patients with no concern of reduced patient satisfaction. Further studies should focus on developing prescribing guidelines for high-risk patients, including those with preoperative opioid needs, and for specific procedures associated with high analgesia requirements.

## Data Availability

All data produced in the present study are available upon reasonable request to the authors

## SUPPLEMENTARY MATERIAL

**Figure S1:**
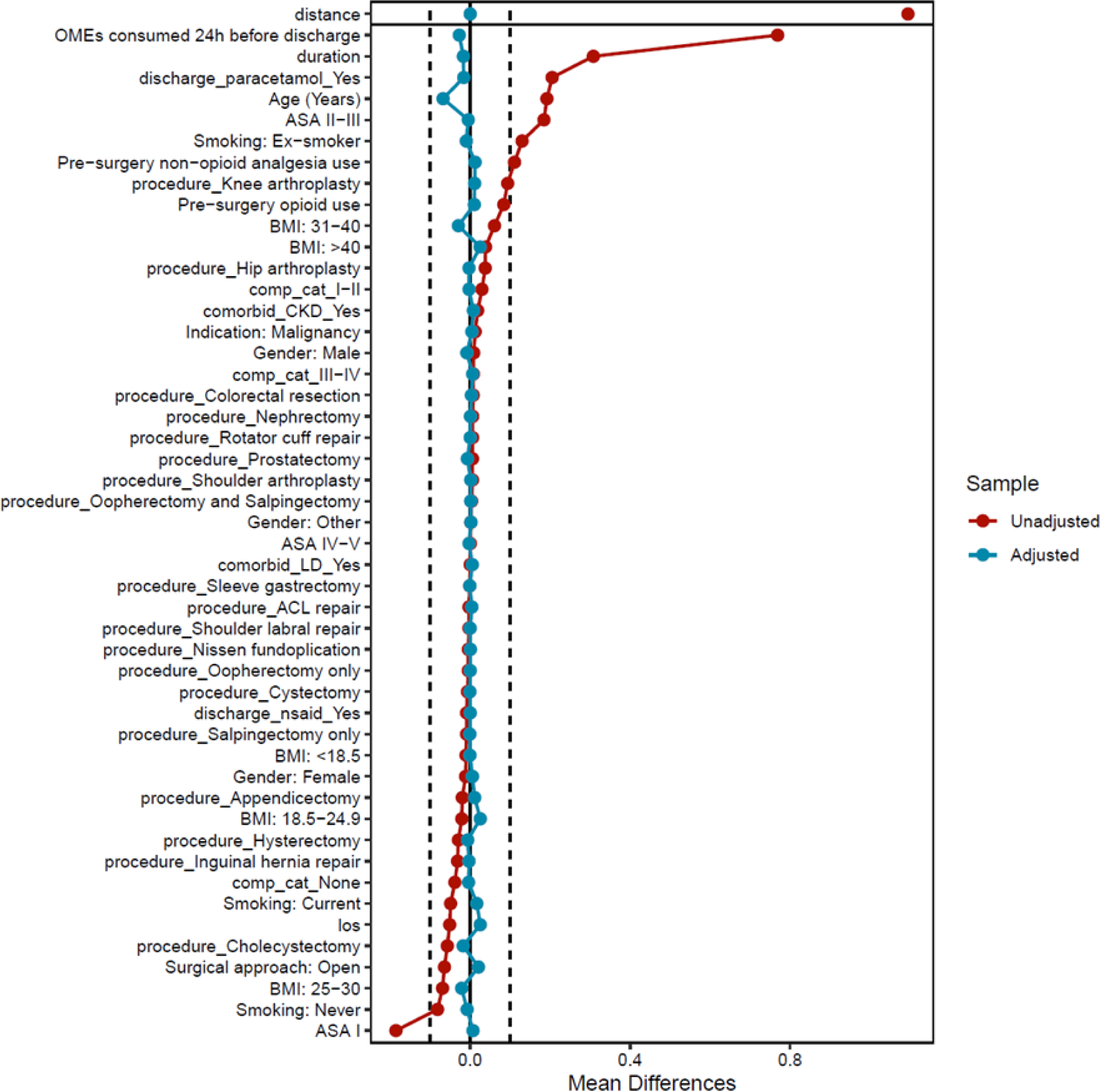
Standardised mean difference of variables between patients discharged with opioid and opioid-free analgesia before and after propensity score matching

**Figure S2.**
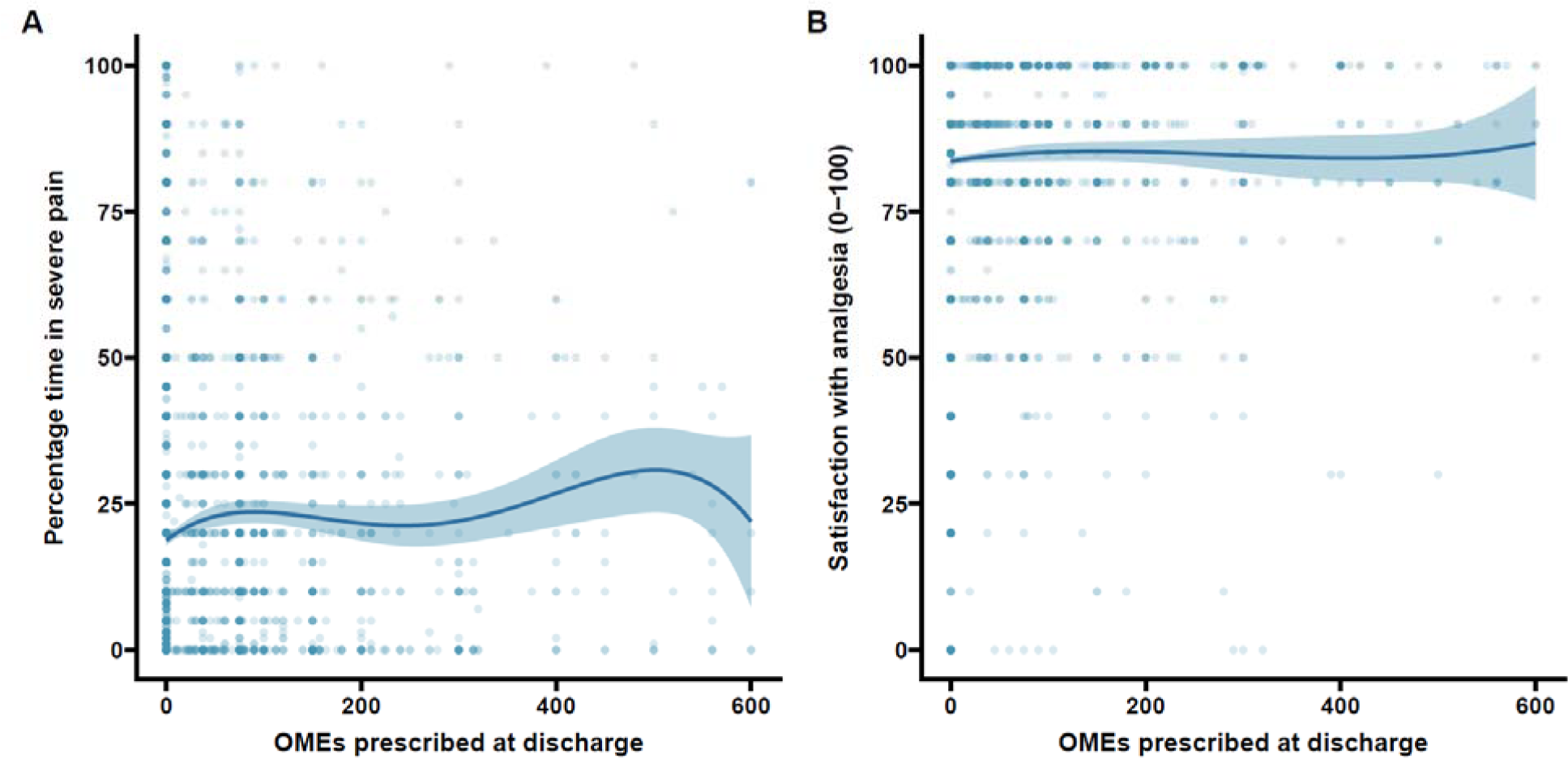
Relationship between quantity of OMEs prescribed at discharge and A) unadjusted time in severe pain and B) unadjusted patient satisfaction with pain treatment adjusted for patient demographics

**Table S1:**
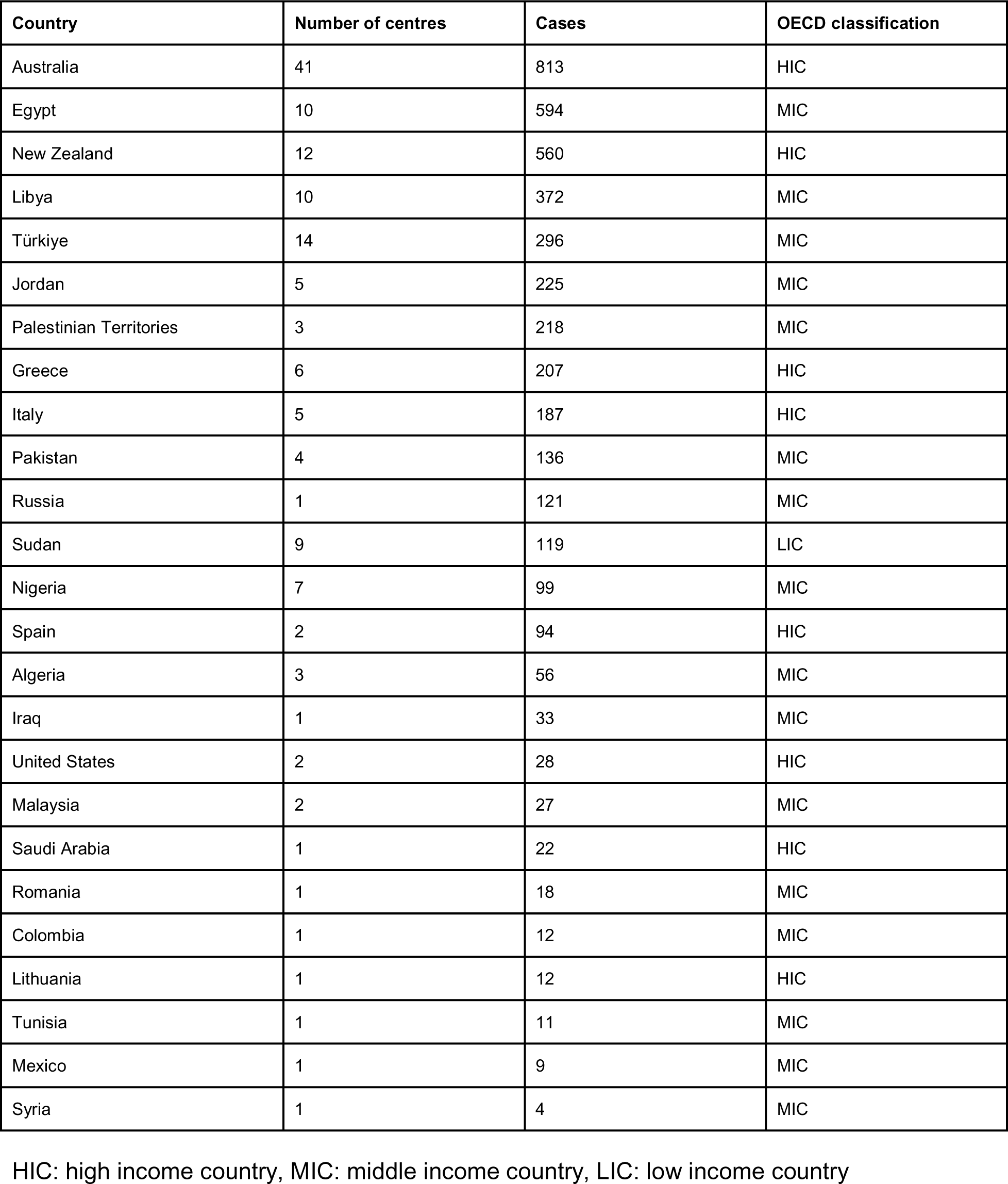
Contributions to study by country.

| Country | Number of centres | Cases | OECD classification |
| --- | --- | --- | --- |
| Australia | 41 | 813 | HIC |
| Egypt | 10 | 594 | MIC |
| New Zealand | 12 | 560 | HIC |
| Libya | 10 | 372 | MIC |
| Türkiye | 14 | 296 | MIC |
| Jordan | 5 | 225 | MIC |
| Palestinian Territories | 3 | 218 | MIC |
| Greece | 6 | 207 | HIC |
| Italy | 5 | 187 | HIC |
| Pakistan | 4 | 136 | MIC |
| Russia | 1 | 121 | MIC |
| Sudan | 9 | 119 | LIC |
| Nigeria | 7 | 99 | MIC |
| Spain | 2 | 94 | HIC |
| Algeria | 3 | 56 | MIC |
| Iraq | 1 | 33 | MIC |
| United States | 2 | 28 | HIC |
| Malaysia | 2 | 27 | MIC |
| Saudi Arabia | 1 | 22 | HIC |
| Romania | 1 | 18 | MIC |
| Colombia | 1 | 12 | MIC |
| Lithuania | 1 | 12 | HIC |
| Tunisia | 1 | 11 | MIC |
| Mexico | 1 | 9 | MIC |
| Syria | 1 | 4 | MIC |

HIC: high income country, MIC: middle income country, LIC: low income country

**Table S2:** Balance diagnostics.

| Variable | Type | Mean SMD unadj | Mean SMD adj |
| --- | --- | --- | --- |
| distance | Distance | 1.007616832 | 0.00017236 |
| Age (Years) | Contin. | 0.149058 | -0.050139333 |
| Sex: Female | Binary | -0.00524441 | 0.009990547 |
| Sex: Male | Binary | 0.002956081 | -0.012278876 |
| Sex: Other | Binary | 0.00228833 | 0.00228833 |
| ASA I | Binary | -0.169717824 | 0.002463279 |
| ASA II-III | Binary | 0.166412571 | -0.004285986 |
| ASA IV-V | Binary | 0.003305253 | 0.001822707 |
| BMI: 18.5-24.9 | Binary | -0.024395843 | 0.010543382 |
| BMI: 25-30 | Binary | -0.068929914 | -0.003954364 |
| BMI: 31-40 | Binary | 0.064590509 | -0.01080799 |
| BMI: >40 | Binary | 0.037283805 | 0.003637039 |
| BMI: <18.5 | Binary | -0.008548556 | 0.000581933 |
| comorbid_CKD_Yes | Binary | 0.016256513 | -0.003281365 |
| comorbid_LD_Yes | Binary | 0.003802037 | 0.004374859 |
| Smoking: Never | Binary | -0.070994432 | 0.002851856 |
| Smoking: Current | Binary | -0.049177788 | -0.002056171 |
| Smoking: Ex-smoker | Binary | 0.12017222 | -0.000795684 |
| procedure_Appendicectomy | Binary | 0.007596863 | 0.007001575 |
| procedure_Cholecystectomy | Binary | -0.033175885 | 0.020255929 |
| procedure_Colorectal resection | Binary | 0.001566499 | -0.014520139 |
| procedure_Inguinal hernia repair | Binary | -0.044810673 | 0.009403452 |
| procedure_Nissen fundoplication | Binary | -0.003951141 | 5.27E-05 |
| procedure_Sleeve gastrectomy | Binary | -0.00052457 | 0.00089529 |
| procedure_ACL repair | Binary | -0.003225448 | -0.000448776 |
| procedure_Knee arthroplasty | Binary | 0.074991077 | -0.002822054 |
| procedure_Hip arthroplasty | Binary | 0.032275593 | 0.001255537 |
| procedure_Rotator cuff repair | Binary | 0.004676835 | 0.001749313 |
| procedure_Shoulder arthroplasty | Binary | 0.005352054 | -0.00361996 |
| procedure_Shoulder labral repair | Binary | -0.00328854 | -1.55E-05 |
| procedure_Cystectomy | Binary | -0.006401966 | -0.001238054 |
| procedure_Nephrectomy | Binary | 0.008053704 | -0.007387922 |
| procedure_Prostatectomy | Binary | 0.004414936 | -0.004025605 |
| procedure_Hysterectomy | Binary | -0.031971203 | -0.008389155 |
| procedure_Oophorectomy and Salpingectomy | Binary | 0.000463022 | 0.001912783 |
| procedure_Oophorectomy only | Binary | -0.004801475 | 0.00088914 |
| procedure_Salpingectomy only | Binary | -0.007239681 | -0.000948612 |
| Surgical approach: Open | Binary | -0.119120762 | 0.02417267 |
| Indication: Malignancy | Binary | 0.005709132 | -0.021959313 |
| Urgency: Emergency | Binary | 0.084354017 | -0.001681727 |
| duration | Contin. | 0.219513659 | -0.081228276 |
| comp_cat_None | Binary | -0.045320384 | 0.017599561 |
| comp_cat_I-II | Binary | 0.037188419 | -0.021473516 |
| comp_cat_III-IV | Binary | 0.008131965 | 0.003873954 |
| OMEs consumed 24h<br>before discharge | Contin. | 0.323538167 | 0.020525119 |
| Pre-surgery opioid use | Binary | 0.082030407 | 0.025397245 |
| Pre-surgery non-opioid<br>analgesia use | Binary | 0.097242416 | 0.005469069 |
| los | Contin. | -0.067801416 | -0.056134528 |
| discharge_paracetamol_<br>Yes | Binary | 0.180030784 | -0.01173816 |
| discharge_nsaid_Yes | Binary | -0.017167751 | -0.010622419 |

**Table S3:** Negative binomial regression model of factors associated with percent time in severe pain post-discharge.

| Variable | Risk ratio | 95% CI | p-value |
| --- | --- | --- | --- |
| Intercept | 23.45 | 15.97 - 34.44 | 0 |
| Age (Years) | 0.99 | 0.99 - 1 | 0.011 |
| Sex: Female | Ref |  |  |
| Sex: Male | 0.87 | 0.77 - 0.99 | 0.034 |
| Sex: Other | 0.99 | 0.18 - 5.36 | 0.994 |
| ASA I | Ref |  |  |
| ASA II-III | 1.01 | 0.86 - 1.18 | 0.914 |
| ASA IV-V | 1.63 | 0.96 - 2.8 | 0.075 |
| BMI: 20-24 | Ref |  |  |
| BMI: 25-30 | 1.07 | 0.92 - 1.25 | 0.358 |
| BMI: 31-40 | 1.18 | 1.02 - 1.38 | 0.03 |
| BMI: >40 | 1.13 | 0.88 - 1.47 | 0.338 |
| BMI: <18.5 | 1.84 | 1.03 - 3.28 | 0.042 |
| Smoking: Never | Ref |  |  |
| Smoking: Current | 1.04 | 0.88 - 1.23 | 0.637 |
| Smoking: Ex-smoker | 1.13 | 0.98 - 1.29 | 0.095 |
| Surgical approach: MIS | Ref |  |  |
| Surgical approach: Open | 1.08 | 0.92 - 1.26 | 0.351 |
| Indication: Benign | Ref |  |  |
| Indication: Malignancy | 0.73 | 0.56 - 0.95 | 0.025 |
| Urgency: Elective |  |  |  |
| Urgency: Emergency | 0.81 | 0.67 - 0.97 | 0.027 |
| Pre-surgery opioid use: No | Ref |  |  |
| Pre-surgery opioid use: Yes | 1.14 | 0.88 - 1.49 | 0.321 |
| Pre-surgery non-opioid analgesia use: No | Ref |  |  |
| Pre-surgery non-opioid analgesia use: Yes | 1.07 | 0.96 - 1.21 | 0.225 |
| Procedure: Appendectomy | Ref |  |  |
| Procedure: Cholecystectomy | 0.71 | 0.58 - 0.86 | 0.001 |
| Procedure: Colorectal resection | 0.91 | 0.62 - 1.33 | 0.628 |
| Procedure: Inguinal hernia repair | 0.83 | 0.62 - 1.11 | 0.209 |
| Procedure: Nissen fundoplication | 0.6 | 0.28 - 1.3 | 0.194 |
| Procedure: Sleeve gastrectomy | 0.68 | 0.42 - 1.11 | 0.128 |
| Procedure: ACL repair | 1.33 | 0.86 - 2.07 | 0.199 |
| Procedure: Knee arthroplasty | 1.4 | 1.05 - 1.88 | 0.023 |
| Procedure: Hip arthroplasty | 1.18 | 0.88 - 1.58 | 0.269 |
| Procedure: Rotator cuff repair | 1.53 | 0.79 - 2.94 | 0.206 |
| Procedure: Shoulder arthroplasty | 1.13 | 0.62 - 2.05 | 0.689 |
| Procedure: Shoulder labral repair | 0.91 | 0.17 - 4.99 | 0.914 |
| Procedure: Cystectomy | 0.72 | 0.32 - 1.61 | 0.423 |
| Procedure: Nephrectomy | 1.24 | 0.82 - 1.87 | 0.304 |
| Procedure: Prostatectomy | 0.88 | 0.57 - 1.38 | 0.591 |
| Procedure: Hysterectomy | 0.89 | 0.62 - 1.27 | 0.517 |
| Procedure: Oophorectomy and salpingectomy | 0.95 | 0.55 - 1.66 | 0.868 |
| Procedure: Oophorectomy only | 1.35 | 0.44 - 4.13 | 0.599 |
| Procedure: Salpingectomy only | 0.94 | 0.43 - 2.02 | 0.868 |
| Clavien Dindo: None | Ref |  |  |
| Clavien Dindo: I-II | 1.24 | 1.06 - 1.45 | 0.01 |
| Clavien Dindo: III-IV | 1 | 0.59 - 1.7 | 0.994 |
| Discharge opioids: No | Ref |  |  |
| Discharge opioids: Yes | 1.52 | 1.31 - 1.76 | 0 |
| OMEs consumed 24h before discharge (per unit) | 1 | 1-1 | 0.143 |

ASA: American Association of Anaesthesiologists Grade, BMI: body mass index ( $\text{kg/m}^2$ ), MIS: minimally invasive surgery. Mixed-effects model with country and hospital centre as random effects.

**Table S4:** Mixed-effects linear regression model of factors associated with satisfaction with pain treatment (0-100)

| Variable | Coefficient | 95% CI | p-value |
| --- | --- | --- | --- |
| Intercept | 82.76 | 78.77 - 86.74 | 0 |
| Age (Years) | 0.04 | -0.02 - 0.09 | 0.191 |
| Sex: Female | Ref |  |  |
| Sex: Male | 2.18 | 0.15 - 4.21 | 0.046 |
| Sex: Other | 2.41 | -18.55 - 23.36 | 0.822 |
| ASA I | Ref |  |  |
| ASA II-III | -1.02 | -2.99 - 0.94 | 0.31 |
| ASA IV-V | -8.17 | -17.78 - 1.44 | 0.11 |
| BMI: 20-24 | Ref |  |  |
| BMI: 25-30 | -1.84 | -3.96 - 0.28 | 0.098 |
| BMI: 31-40 | -1.34 | -3.39 - 0.72 | 0.207 |
| BMI: >40 | -3.01 | -7.25 - 1.23 | 0.178 |
| BMI: <18.5 | -12.22 | -20.7 - -3.74 | 0.009 |
| Smoking: Never | Ref |  |  |
| Smoking: Current | -2.38 | -4.75 - -0.02 | 0.056 |
| Smoking: Ex-smoker | -0.2 | -2.31 - 1.91 | 0.852 |
| Surgical approach: MIS | Ref |  |  |
| Surgical approach: Open | -1.4 | -3.51 - 0.72 | 0.201 |
| Indication: Benign | Ref |  |  |
| Indication: Malignancy | 3.64 | 0.27 - 7 | 0.04 |
| Urgency: Elective | Ref |  |  |
| Urgency: Emergency | 1.25 | -0.86 - 3.36 | 0.248 |
| Pre-surgery opioid use: No | Ref |  |  |
| Pre-surgery opioid use: Yes | -3.76 | -7.19 - -0.33 | 0.041 |
| Pre-surgery non-opioid analgesia use: No | Ref |  |  |
| Pre-surgery non-opioid analgesia use: Yes | -0.87 | -2.38 - 0.65 | 0.262 |
| Procedure: Appendectomy | Ref |  |  |
| Procedure: Cholecystectomy | 3.15 | 0.61 - 5.7 | 0.018 |
| Procedure: Colorectal resection | -2.94 | -6.61 - 0.73 | 0.119 |
| Procedure: Inguinal hernia repair | 1.6 | -1.52 - 4.71 | 0.316 |
| Procedure: Nissen fundoplication | -5.38 | -15.24 - 4.48 | 0.285 |
| Procedure: Sleeve gastrectomy | 3.14 | -2.91 - 9.18 | 0.311 |
| Procedure: ACL repair | -10.01 | -15.56 - -4.47 | 0 |
| Procedure: Knee arthroplasty | -3.27 | -7.46 - 0.92 | 0.131 |
| Procedure: Hip arthroplasty | -2.05 | -6.65 - 2.55 | 0.385 |
| Procedure: Rotator cuff repair | -5.38 | -13.05 - 2.3 | 0.17 |
| Procedure: Shoulder arthroplasty | -0.65 | -12.01 - 10.72 | 0.912 |
| Procedure: Shoulder labral repair | 2.81 | -19.99 - 25.62 | 0.809 |
| Procedure: Cystectomy | -6.37 | -22.17 - 9.43 | 0.438 |
| Procedure: Nephrectomy | -3.34 | -8.64 - 1.97 | 0.223 |
| Procedure: Prostatectomy | -3.62 | -9.33 - 2.08 | 0.218 |
| Procedure: Hysterectomy | 1.09 | -3.88 - 6.06 | 0.67 |
| Procedure: Oophorectomy and salpingectomy | -4.59 | -12.82 - 3.64 | 0.279 |
| Procedure: Oophorectomy only | -3.32 | -19.18 - 12.55 | 0.683 |
| Procedure: Salpingectomy only | 3.3 | -6.78 - 13.39 | 0.522 |
| Clavien Dindo: None | Ref |  |  |
| Clavien Dindo: I-II | -3.62 | -6.47 - -0.78 | 0.023 |
| Clavien Dindo: III-IV | -1.99 | -8.01 - 4.03 | 0.518 |
| Discharge opioids: No | Ref |  |  |
| Discharge opioids: Yes | 0.92 | -1.52 - 3.36 | 0.468 |
| OMEs consumed 24h before discharge (per unit) | 0 | -0.01 - 0 | 0.341 |

ASA: American Association of Anaesthesiologists Grade, BMI: body mass index ( $\text{kg/m}^2$ ), MIS: minimally invasive surgery. Mixed-effects model with country and hospital centre as random effects.

**Table S5:** Mixed-effects linear regression model of factors associated with EQ-5D-5L composite score (0-100)

| Variable | Coefficient | 95% CI | p-value |
| --- | --- | --- | --- |
| Intercept | 86.25 | 82.31 - 90.2 | 0 |
| Age (Years) | 0.03 | -0.02 - 0.07 | 0.286 |
| Sex: Female | Ref |  |  |
| Sex: Male | 4.28 | 2.41 - 6.14 | 0 |
| Sex: Other | 18.31 | -0.11 - 36.73 | 0.051 |
| ASA I | Ref |  |  |
| ASA II-III | -2.01 | -3.52 - -0.5 | 0.01 |
| ASA IV-V | -6.44 | -13.48 - 0.6 | 0.081 |
| BMI: 20-24 | Ref |  |  |
| BMI: 25-30 | -0.98 | -2.54 - 0.58 | 0.222 |
| BMI: 31-40 | -2.44 | -4.73 - -0.15 | 0.048 |
| BMI: >40 | -1.98 | -6.04 - 2.08 | 0.351 |
| BMI: <18.5 | -15.85 | -25.53 - -6.17 | 0.006 |
| Smoking: Never | Ref |  |  |
| Smoking: Current | -0.85 | -3.09 - 1.4 | 0.465 |
| Smoking: Ex-smoker | -0.26 | -2.12 - 1.61 | 0.788 |
| Surgical approach: MIS | Ref |  |  |
| Surgical approach: Open | -1.95 | -3.91 - 0.01 | 0.058 |
| Indication: Benign | Ref |  |  |
| Indication: Malignancy | 1.31 | -1.83 - 4.45 | 0.418 |
| Urgency: Elective | Ref |  |  |
| Urgency: Emergency | -1.06 | -3.03 - 0.92 | 0.299 |
| Pre-surgery opioid use: No | Ref |  |  |
| Pre-surgery opioid use: Yes | -5.03 | -8.21 - -1.84 | 0.005 |
| Pre-surgery non-opioid analgesia use: No | Ref |  |  |
| Pre-surgery non-opioid analgesia use: Yes | -1.32 | -2.86 - 0.23 | 0.099 |
| Procedure: Appendectomy | Ref |  |  |
| Procedure: Cholecystectomy | 4.36 | 1.9 - 6.83 | 0.001 |
| Procedure: Colorectal resection | -1.58 | -5.9 - 2.73 | 0.479 |
| Procedure: Inguinal hernia repair | -1.79 | -5.69 - 2.11 | 0.378 |
| Procedure: Nissen fundoplication | -8.26 | -16.77 - 0.25 | 0.057 |
| Procedure: Sleeve gastrectomy | 0.64 | -6.05 - 7.32 | 0.853 |
| Procedure: ACL repair | -18.44 | -24.62 - -12.27 | 0 |
| Procedure: Knee arthroplasty | -10.96 | -15.79 - -6.12 | 0 |
| Procedure: Hip arthroplasty | -13.3 | -19.16 - -7.44 | 0 |
| Procedure: Rotator cuff repair | -11.49 | -19.56 - -3.41 | 0.007 |
| Procedure: Shoulder arthroplasty | -15.15 | -22.36 - -7.95 | 0 |
| Procedure: Shoulder labral repair | -0.61 | -20.2 - 18.98 | 0.952 |
| Procedure: Cystectomy | 5.01 | -4.91 - 14.92 | 0.325 |
| Procedure: Nephrectomy | -8.54 | -13.95 - -3.14 | 0.004 |
| Procedure: Prostatectomy | -4.89 | -10.95 - 1.17 | 0.126 |
| Procedure: Hysterectomy | -2.3 | -8.88 - 4.28 | 0.503 |
| Procedure: Oophorectomy and salpingectomy | -4.19 | -11.07 - 2.69 | 0.236 |
| Procedure: Oophorectomy only | -7.01 | -21 - 6.98 | 0.328 |
| Procedure: Salpingectomy only | 1.8 | -7.12 - 10.72 | 0.693 |
| Clavien Dindo: None | Ref |  |  |
| Clavien Dindo: I-II | -1.72 | -3.68 - 0.25 | 0.096 |
| Clavien Dindo: III-IV | -10.21 | -17.7 - -2.72 | 0.014 |
| Discharge opioids: No | Ref |  |  |
| Discharge opioids: Yes | -2.27 | -3.82 - -0.72 | 0.005 |
| OMEs consumed 24h before discharge (per unit) | 0 | -0.01 - 0 | 0.44 |

ASA: American Association of Anaesthesiologists Grade, BMI: body mass index ( $\text{kg/m}^2$ ), MIS: minimally invasive surgery. Mixed-effects model with country and hospital centre as random effects.

**Table S6:** Mixed-effects linear regression model of factors associated with EQ-VAS QoL score (0-100)

| Variable | Coefficient | 95% CI | p-value |
| --- | --- | --- | --- |
| Intercept | 87.59 | 83.72 - 91.46 | 0 |
| Age (Years) | -0.08 | -0.14 - -0.03 | 0.006 |
| Sex: Female | Ref |  |  |
| Sex: Male | 1.7 | 0.16 - 3.24 | 0.034 |
| Sex: Other | 6.13 | -12.8 - 25.06 | 0.526 |
| ASA I | Ref |  |  |
| ASA II-III | -0.57 | -2.28 - 1.13 | 0.512 |
| ASA IV-V | -5.17 | -11.48 - 1.13 | 0.111 |
| BMI: 20-24 | Ref |  |  |
| BMI: 25-30 | -1.19 | -2.67 - 0.28 | 0.113 |
| BMI: 31-40 | -2.82 | -4.95 - -0.7 | 0.014 |
| BMI: >40 | -5.26 | -9.04 - -1.48 | 0.012 |
| BMI: <18.5 | -12.54 | -20.28 - -4.8 | 0.004 |
| Smoking: Never | Ref |  |  |
| Smoking: Current | -1.19 | -2.67 - 0.28 | 0.113 |
| Smoking: Ex-smoker | -2.82 | -4.95 - -0.7 | 0.014 |
| Surgical approach: MIS | Ref |  |  |
| Surgical approach: Open | -1.63 | -3.31 - 0.05 | 0.058 |
| Indication: Benign | Ref |  |  |
| Indication: Malignancy | 0.58 | -3.14 - 4.29 | 0.764 |
| Urgency: Elective | Ref |  |  |
| Urgency: Emergency | -1.35 | -3 - 0.3 | 0.109 |
| Pre-surgery opioid use: No | Ref |  |  |
| Pre-surgery opioid use: Yes | -2.59 | -5.84 - 0.65 | 0.131 |
| Pre-surgery non-opioid analgesia use: No | Ref |  |  |
| Pre-surgery non-opioid analgesia use: Yes | -0.83 | -2.41 - 0.76 | 0.311 |
| Procedure: Appendectomy | Ref |  |  |
| Procedure: Cholecystectomy | 3.56 | 1.37 - 5.75 | 0.002 |
| Procedure: Colorectal resection | -3.71 | -7.55 - 0.12 | 0.065 |
| Procedure: Inguinal hernia repair | 1.25 | -1.92 - 4.42 | 0.441 |
| Procedure: Nissen fundoplication | -3.23 | -12.36 - 5.9 | 0.488 |
| Procedure: Sleeve gastrectomy | 2.98 | -3.46 - 9.41 | 0.37 |
| Procedure: ACL repair | -5.57 | -10.62 - -<br>0.53 | 0.031 |
| Procedure: Knee arthroplasty | -0.92 | -4.74 - 2.9 | 0.638 |
| Procedure: Hip arthroplasty | -1.19 | -6.14 - 3.75 | 0.64 |
| Procedure: Rotator cuff repair | 0.52 | -6.53 - 7.57 | 0.885 |
| Procedure: Shoulder arthroplasty | 1.95 | -6.17 - 10.08 | 0.64 |
| Procedure: Shoulder labral repair | -10.26 | -29.45 - 8.93 | 0.295 |
| Procedure: Cystectomy | 0.36 | -10.22 -<br>10.93 | 0.948 |
| Procedure: Nephrectomy | -6.44 | -11.08 - -1.8 | 0.008 |
| Procedure: Prostatectomy | 0.78 | -4.4 - 5.96 | 0.768 |
| Procedure: Hysterectomy | -3.05 | -7.6 - 1.5 | 0.196 |
| Procedure: Oophorectomy and salpingectomy | -3.27 | -9.72 - 3.17 | 0.321 |
| Procedure: Oophorectomy only | -0.65 | -14.38 -<br>13.08 | 0.926 |
| Procedure: Salpingectomy only | -4.25 | -12.67 - 4.16 | 0.322 |
| Clavien Dindo: None | Ref |  |  |
| Clavien Dindo: I-II | -1.1 | -3.17 - 0.96 | 0.303 |
| Clavien Dindo: III-IV | -10.38 | -16.89 - -<br>3.86 | 0.003 |
| Discharge opioids: No | Ref |  |  |
| Discharge opioids: Yes | -2.82 | -4.51 - -1.14 | 0.002 |
| OMEs consumed 24h before discharge (per unit) | 0 | -0.01 - 0.01 | 0.758 |

ASA: American Association of Anaesthesiologists Grade, BMI: body mass index ( $\text{kg/m}^2$ ), MIS: minimally invasive surgery. Mixed-effects model with country and hospital centre as random effects.

**Table S7:** Mixed-effects binary logistic regression model of factors associated with healthcare presentations for further pain relief.

| Variable | Odds ratio | 95% CI | p-value |
| --- | --- | --- | --- |
| Intercept | 0.09 | 0.05 - 0.19 | 0 |
| Age (Years) | 0.99 | 0.98 - 1 | 0.065 |
| Sex: Female | Ref |  |  |
| Sex: Male | 1.03 | 0.77 - 1.38 | 0.825 |
| Sex: Other | 2.05 | 0.15 - 28.23 | 0.59 |
| ASA I | Ref |  |  |
| ASA II-III | 1.28 | 0.92 - 1.78 | 0.138 |
| ASA IV-V | 3.65 | 1.29 - 10.32 | 0.016 |
| BMI: 20-24 | Ref |  |  |
| BMI: 25-30 | 1.3 | 0.82 - 2.08 | 0.275 |
| BMI: 31-40 | 1.29 | 0.82 - 2.03 | 0.288 |
| BMI: >40 | 1.33 | 0.62 - 2.84 | 0.469 |
| BMI: <18.5 | 3.5 | 1.1 - 11.11 | 0.04 |
| Smoking: Never | Ref |  |  |
| Smoking: Current | 1.39 | 0.93 - 2.08 | 0.113 |
| Smoking: Ex-smoker | 0.97 | 0.62 - 1.51 | 0.897 |
| Surgical approach: MIS | Ref |  |  |
| Surgical approach: Open | 1.46 | 1.03 - 2.09 | 0.037 |
| Indication: Benign | Ref |  |  |
| Indication: Malignancy | 0.4 | 0.2 - 0.81 | 0.014 |
| Urgency: Elective | Ref |  |  |
| Urgency: Emergency | 0.72 | 0.44 - 1.19 | 0.21 |
| Pre-surgery opioid use: No | Ref |  |  |
| Pre-surgery opioid use: Yes | 2.52 | 1.62 - 3.91 | 0 |
| Pre-surgery non-opioid analgesia use: No | Ref |  |  |
| Pre-surgery non-opioid analgesia use: Yes | 1.08 | 0.79 - 1.46 | 0.632 |
| Procedure: Appendectomy | Ref |  |  |
| Procedure: Cholecystectomy | 0.72 | 0.47 - 1.11 | 0.143 |
| Procedure: Colorectal resection | 0.84 | 0.41 - 1.71 | 0.624 |
| Procedure: Inguinal hernia repair | 0.55 | 0.27 - 1.1 | 0.094 |
| Procedure: Nissen fundoplication | 1.11 | 0.18 - 6.96 | 0.913 |
| Procedure: Sleeve gastrectomy | 0.92 | 0.26 - 3.31 | 0.902 |
| Procedure: ACL repair | 0.96 | 0.33 - 2.81 | 0.942 |
| Procedure: Knee arthroplasty | 1.3 | 0.55 - 3.06 | 0.556 |
| Procedure: Hip arthroplasty | 0.68 | 0.27 - 1.7 | 0.411 |
| Procedure: Rotator cuff repair | 0.2 | 0.02 - 1.74 | 0.145 |
| Procedure: Shoulder arthroplasty | 1.26 | 0.31 - 5.16 | 0.754 |
| Procedure: Shoulder labral repair | 3.26 | 0.08 - 137.67 | 0.537 |
| Procedure: Cystectomy | 0.04 | 0 - 154.37 | 0.435 |
| Procedure: Nephrectomy | 1.18 | 0.43 - 3.19 | 0.752 |
| Procedure: Prostatectomy | 0.67 | 0.18 - 2.49 | 0.557 |
| Procedure: Hysterectomy | 0.66 | 0.25 - 1.75 | 0.41 |
| Procedure: Oophorectomy and salpingectomy | 0.59 | 0.09 - 3.98 | 0.591 |
| Procedure: Oophorectomy only | 1.55 | 0.19 - 12.65 | 0.685 |
| Procedure: Salpingectomy only | 0.83 | 0.18 - 3.77 | 0.806 |
| Clavien Dindo: None | Ref |  |  |
| Clavien Dindo: I-II | 2.03 | 1.45 - 2.85 | 0 |
| Clavien Dindo: III-IV | 0.58 | 0.1 - 3.24 | 0.536 |
| Discharge opioids: No | Ref |  |  |
| Discharge opioids: Yes | 1.01 | 0.7 - 1.46 | 0.948 |
| OMEs consumed 24h before discharge (per unit) | 1 | 1-1 | 0.004 |

ASA: American Association of Anaesthesiologists Grade, BMI: body mass index ( $\text{kg/m}^2$ ), MIS: minimally invasive surgery. Mixed-effects model with country and hospital centre as random effects.

**Table S8:**
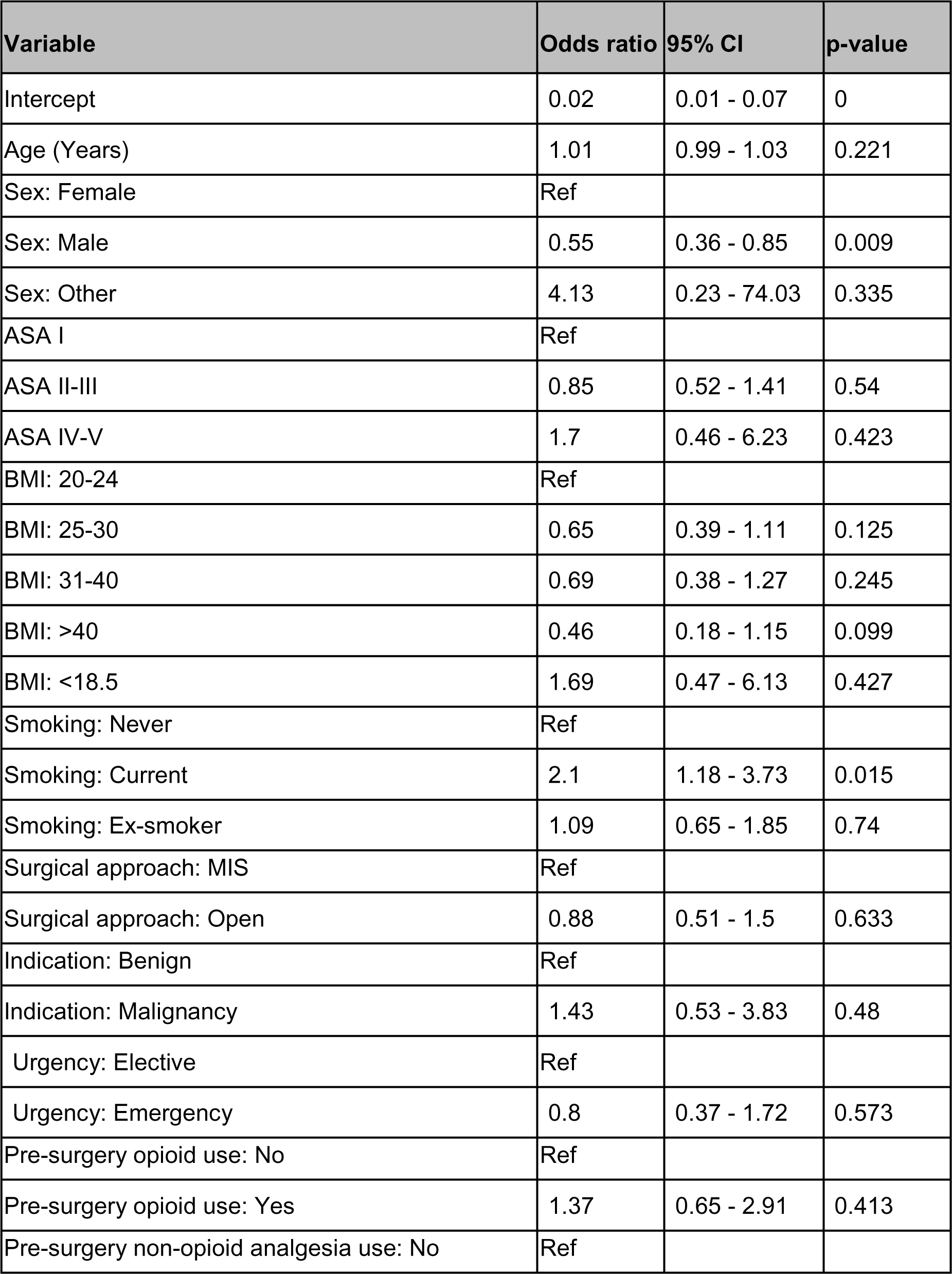
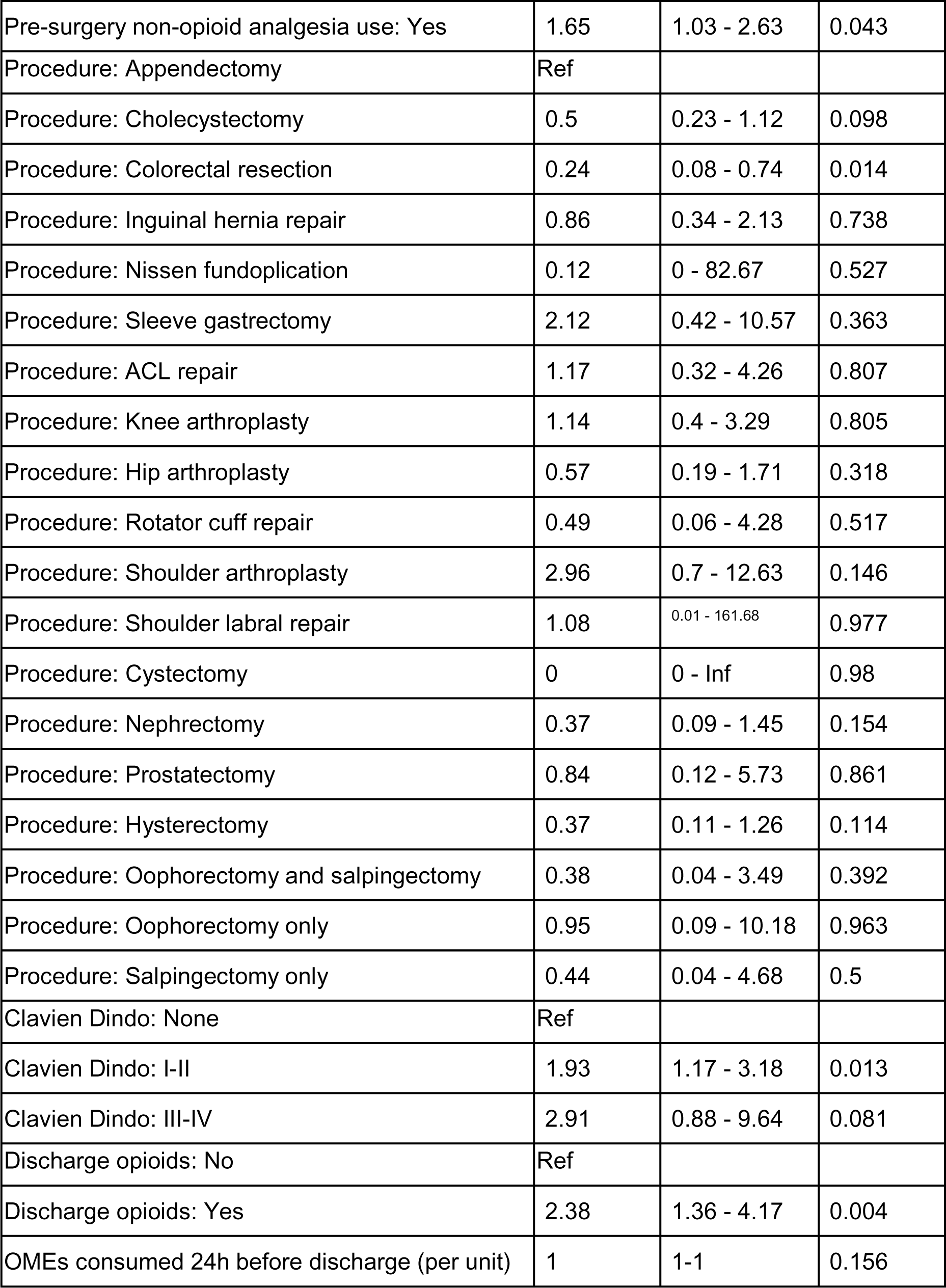
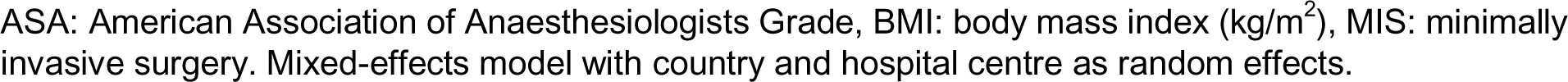
Mixed-effects binary logistic regression model of factors associated with healthcare presentations for side effects of analgesia.

| Variable | Odds ratio | 95% CI | p-value |
| --- | --- | --- | --- |
| Intercept | 0.02 | 0.01 - 0.07 | 0 |
| Age (Years) | 1.01 | 0.99 - 1.03 | 0.221 |
| Sex: Female | Ref |  |  |
| Sex: Male | 0.55 | 0.36 - 0.85 | 0.009 |
| Sex: Other | 4.13 | 0.23 - 74.03 | 0.335 |
| ASA I | Ref |  |  |
| ASA II-III | 0.85 | 0.52 - 1.41 | 0.54 |
| ASA IV-V | 1.7 | 0.46 - 6.23 | 0.423 |
| BMI: 20-24 | Ref |  |  |
| BMI: 25-30 | 0.65 | 0.39 - 1.11 | 0.125 |
| BMI: 31-40 | 0.69 | 0.38 - 1.27 | 0.245 |
| BMI: >40 | 0.46 | 0.18 - 1.15 | 0.099 |
| BMI: <18.5 | 1.69 | 0.47 - 6.13 | 0.427 |
| Smoking: Never | Ref |  |  |
| Smoking: Current | 2.1 | 1.18 - 3.73 | 0.015 |
| Smoking: Ex-smoker | 1.09 | 0.65 - 1.85 | 0.74 |
| Surgical approach: MIS | Ref |  |  |
| Surgical approach: Open | 0.88 | 0.51 - 1.5 | 0.633 |
| Indication: Benign | Ref |  |  |
| Indication: Malignancy | 1.43 | 0.53 - 3.83 | 0.48 |
| Urgency: Elective | Ref |  |  |
| Urgency: Emergency | 0.8 | 0.37 - 1.72 | 0.573 |
| Pre-surgery opioid use: No | Ref |  |  |
| Pre-surgery opioid use: Yes | 1.37 | 0.65 - 2.91 | 0.413 |
| Pre-surgery non-opioid analgesia use: No | Ref |  |  |
| Pre-surgery non-opioid analgesia use: Yes | 1.65 | 1.03 - 2.63 | 0.043 |
| Procedure: Appendectomy | Ref |  |  |
| Procedure: Cholecystectomy | 0.5 | 0.23 - 1.12 | 0.098 |
| Procedure: Colorectal resection | 0.24 | 0.08 - 0.74 | 0.014 |
| Procedure: Inguinal hernia repair | 0.86 | 0.34 - 2.13 | 0.738 |
| Procedure: Nissen fundoplication | 0.12 | 0 - 82.67 | 0.527 |
| Procedure: Sleeve gastrectomy | 2.12 | 0.42 - 10.57 | 0.363 |
| Procedure: ACL repair | 1.17 | 0.32 - 4.26 | 0.807 |
| Procedure: Knee arthroplasty | 1.14 | 0.4 - 3.29 | 0.805 |
| Procedure: Hip arthroplasty | 0.57 | 0.19 - 1.71 | 0.318 |
| Procedure: Rotator cuff repair | 0.49 | 0.06 - 4.28 | 0.517 |
| Procedure: Shoulder arthroplasty | 2.96 | 0.7 - 12.63 | 0.146 |
| Procedure: Shoulder labral repair | 1.08 | 0.01 - 161.68 | 0.977 |
| Procedure: Cystectomy | 0 | 0 - Inf | 0.98 |
| Procedure: Nephrectomy | 0.37 | 0.09 - 1.45 | 0.154 |
| Procedure: Prostatectomy | 0.84 | 0.12 - 5.73 | 0.861 |
| Procedure: Hysterectomy | 0.37 | 0.11 - 1.26 | 0.114 |
| Procedure: Oophorectomy and salpingectomy | 0.38 | 0.04 - 3.49 | 0.392 |
| Procedure: Oophorectomy only | 0.95 | 0.09 - 10.18 | 0.963 |
| Procedure: Salpingectomy only | 0.44 | 0.04 - 4.68 | 0.5 |
| Clavien Dindo: None | Ref |  |  |
| Clavien Dindo: I-II | 1.93 | 1.17 - 3.18 | 0.013 |
| Clavien Dindo: III-IV | 2.91 | 0.88 - 9.64 | 0.081 |
| Discharge opioids: No | Ref |  |  |
| Discharge opioids: Yes | 2.38 | 1.36 - 4.17 | 0.004 |
| OMEs consumed 24h before discharge (per unit) | 1 | 1-1 | 0.156 |

ASA: American Association of Anaesthesiologists Grade, BMI: body mass index ( $\text{kg/m}^2$ ), MIS: minimally invasive surgery. Mixed-effects model with country and hospital centre as random effects.

**Table S9:** Mixed-effects binary logistic regression model of factors associated with patients reporting ‘too little analgesia’ prescribed at 7 days.

| Variable | Odds ratio | 95% CI | p-value |
| --- | --- | --- | --- |
| Intercept | 0.29 | 0.14 – 0.63 | 0.002 |
| Age (Years) | 0.99 | 0.99 – 1.00 | 0.096 |
| Sex: Female | Ref |  |  |
| Sex: Male | 0.8 | 0.66 – 0.97 | 0.023 |
| ASA I | Ref |  |  |
| ASA II-III | 1.12 | 0.90 – 1.38 | 0.313 |
| ASA IV-V | 2.66 | 1.27 – 5.54 | 0.009 |
| BMI: 20-24 | Ref |  |  |
| BMI: 25-30 | 1.11 | 0.91 – 1.37 | 0.301 |
| BMI: 31-40 | 1.23 | 0.97 – 1.56 | 0.091 |
| BMI: >40 | 1.35 | 0.89 – 2.06 | 0.161 |
| BMI: <18.5 | 1.63 | 0.90 – 2.97 | 0.11 |
| Smoking: Never | Ref |  |  |
| Smoking: Current | 1.22 | 0.96 – 1.55 | 0.101 |
| Smoking: Ex-smoker | 1.25 | 0.99 – 1.60 | 0.066 |
| Surgical approach: MIS | Ref |  |  |
| Surgical approach: Open | 0.87 | 0.69 – 1.09 | 0.215 |
| Indication: Benign | Ref |  |  |
| Indication: Malignancy | 0.76 | 0.52 – 1.09 | 0.136 |
| Urgency: Elective | Ref |  |  |
| Urgency: Emergency | 1.16 | 0.90 – 1.50 | 0.251 |
| Pre-surgery opioid use: No | Ref |  |  |
| Pre-surgery opioid use: Yes | 1.94 | 1.34 – 2.82 | <0.001 |
| Pre-surgery non-opioid analgesia use: No | Ref |  |  |
| Pre-surgery non-opioid analgesia use: Yes | 1.09 | 0.89 – 1.34 | 0.412 |
| Procedure: Appendectomy | Ref |  |  |
| Procedure: Cholecystectomy | 0.79 | 0.58 – 1.08 | 0.139 |
| Procedure: Colorectal resection | 1.3 | 0.82 – 2.06 | 0.268 |
| Procedure: Inguinal hernia repair | 1.15 | 0.78 – 1.71 | 0.483 |
| Procedure: Nissen fundoplication | 1.21 | 0.46 – 3.16 | 0.696 |
| Procedure: Sleeve gastrectomy | 1.41 | 0.71 – 2.80 | 0.326 |
| Procedure: ACL repair | 2.83 | 1.53 – 5.24 | 0.001 |
| Procedure: Knee arthroplasty | 2.37 | 1.43 – 3.93 | 0.001 |
| Procedure: Hip arthroplasty | 1.63 | 0.95 – 2.78 | 0.076 |
| Procedure: Rotator cuff repair | 1.04 | 0.31 – 3.52 | 0.952 |
| Procedure: Shoulder arthroplasty | 4.39 | 1.50 – 12.91 | 0.007 |
| Procedure: Shoulder labral repair | 2.09 | 0.58 – 7.56 | 0.26 |
| Procedure: Cystectomy | 3.06 | 1.22 – 7.68 | 0.018 |
| Procedure: Nephrectomy | 1.47 | 0.76 – 2.85 | 0.258 |
| Procedure: Prostatectomy | 1.45 | 0.74 – 2.83 | 0.276 |
| Procedure: Hysterectomy | 0.9 | 0.56 – 1.47 | 0.685 |
| Procedure: Oophorectomy and salpingectomy | 1.53 | 0.68 – 3.47 | 0.307 |
| Procedure: Oophorectomy only | 1.35 | 0.45 – 4.02 | 0.595 |
| Procedure: Salpingectomy only | 0.83 | 0.35 – 1.93 | 0.658 |
| Clavien Dindo: None | Ref |  |  |
| Clavien Dindo: I-II | 1.41 | 1.13 – 1.76 | 0.003 |
| Clavien Dindo: III-IV | 1.28 | 0.55 – 2.99 | 0.572 |
| Discharge paracetamol: No | Ref |  |  |
| Discharge paracetamol: Yes | 0.65 | 0.52 – 0.81 | <0.001 |
| Discharge NSAIDs: No | Ref |  |  |
| Discharge NSAIDs: Yes | 0.91 | 0.75 – 1.09 | 0.307 |
| Discharge opioids: No | Ref |  |  |
| Discharge opioids: Yes | 0.59 | 0.46 – 0.75 | <0.001 |
| OMEs consumed 24h before discharge (per unit) | 1 | 1-1 | 0.156 |

ASA: American Association of Anaesthesiologists Grade, BMI: body mass index ( $\text{kg/m}^2$ ), MIS: minimally invasive surgery. Mixed-effects model with country and hospital centre as random effects.

**Table S10:** Binary logistic regression model for risk of opioid prescription between HIC and LMIC.

| Variable | Odds ratio | 95% CI | p-value |
| --- | --- | --- | --- |
| Intercept | 2.16 | 1.36 - 3.43 | 0.002 |
| Age (Years) | 0.99 | 0.98 - 0.99 | 0 |
| Sex: Female | Ref |  |  |
| Sex: Male | 0.95 | 0.78 - 1.17 | 0.63 |
| ASA I | Ref |  |  |
| ASA II-III | 0.81 | 0.65 - 1.01 | 0.058 |
| ASA IV-V | 1.82 | 0.81 - 4.1 | 0.151 |
| BMI: 20-24 | Ref |  |  |
| BMI: 25-30 | 1.04 | 0.83 - 1.31 | 0.738 |
| BMI: 31-40 | 0.91 | 0.73 - 1.14 | 0.414 |
| BMI: >40 | 0.8 | 0.51 - 1.26 | 0.338 |
| BMI: <18.5 | 1.47 | 0.7 - 3.07 | 0.312 |
| Smoking: Never | Ref |  |  |
| Smoking: Current | 1.1 | 0.83 - 1.44 | 0.506 |
| Smoking: Ex-smoker | 0.76 | 0.62 - 0.94 | 0.011 |
| Surgical approach: MIS | Ref |  |  |
| Surgical approach: Open | 1.98 | 1.56 - 2.52 | 0 |
| Indication: Benign | Ref |  |  |
| Indication: Malignancy | 1.28 | 0.87 - 1.87 | 0.214 |
| Urgency: Elective | Ref |  |  |
| Urgency: Emergency | 0.62 | 0.48 - 0.79 | 0 |
| Pre-surgery opioid use: No | Ref |  |  |
| Pre-surgery opioid use: Yes | 0.83 | 0.61 - 1.13 | 0.249 |
| Pre-surgery non-opioid analgesia use: No | Ref |  |  |
| Pre-surgery non-opioid analgesia use: Yes | 1.12 | 0.9 - 1.39 | 0.316 |
| Procedure: Appendectomy | Ref |  |  |
| Procedure: Cholecystectomy | 1.5 | 1.05 - 2.13 | 0.031 |
| Procedure: Colorectal resection | 0.76 | 0.48 - 1.21 | 0.246 |
| Procedure: Inguinal hernia repair | 1.1 | 0.72 - 1.67 | 0.656 |
| Procedure: Nissen fundoplication | 1.43 | 0.38 - 5.46 | 0.599 |
| Procedure: Sleeve gastrectomy | 1.5 | 0.7 - 3.22 | 0.304 |
| Procedure: ACL repair | 1.82 | 0.86 - 3.83 | 0.118 |
| Procedure: Knee arthroplasty | 0.93 | 0.55 - 1.59 | 0.793 |
| Procedure: Hip arthroplasty | 0.74 | 0.38 - 1.44 | 0.38 |
| Procedure: Rotator cuff repair | 1.06 | 0.35 - 3.26 | 0.918 |
| Procedure: Shoulder arthroplasty | 0.61 | 0.13 - 3 | 0.554 |
| Procedure: Shoulder labral repair | 2.32 | 0.17 - 31.49 | 0.527 |
| Procedure: Cystectomy | 0.55 | 0.14 - 2.18 | 0.395 |
| Procedure: Nephrectomy | 0.99 | 0.48 - 2.05 | 0.986 |
| Procedure: Prostatectomy | 0.74 | 0.38 - 1.46 | 0.393 |
| Procedure: Hysterectomy | 0.86 | 0.48 - 1.56 | 0.621 |
| Procedure: Oophorectomy and salpingectomy | 1.29 | 0.44 - 3.78 | 0.64 |
| Procedure: Oophorectomy only | 2.39 | 0.46 - 12.33 | 0.298 |
| Procedure: Salpingectomy only | 0.55 | 0.1 - 3.01 | 0.499 |
| Clavien Dindo: None | Ref |  |  |
| Clavien Dindo: I-II | 0.89 | 0.71 - 1.12 | 0.324 |
| Clavien Dindo: III-IV | 1.12 | 0.53 - 2.36 | 0.769 |
| OMEs consumed 24h before discharge (per unit) | 1 | 1-Jan | 0 |
| LMIC | Ref |  |  |
| HIC | 9.1 | 7.7-11.1 | 0 |

**Table S11:**
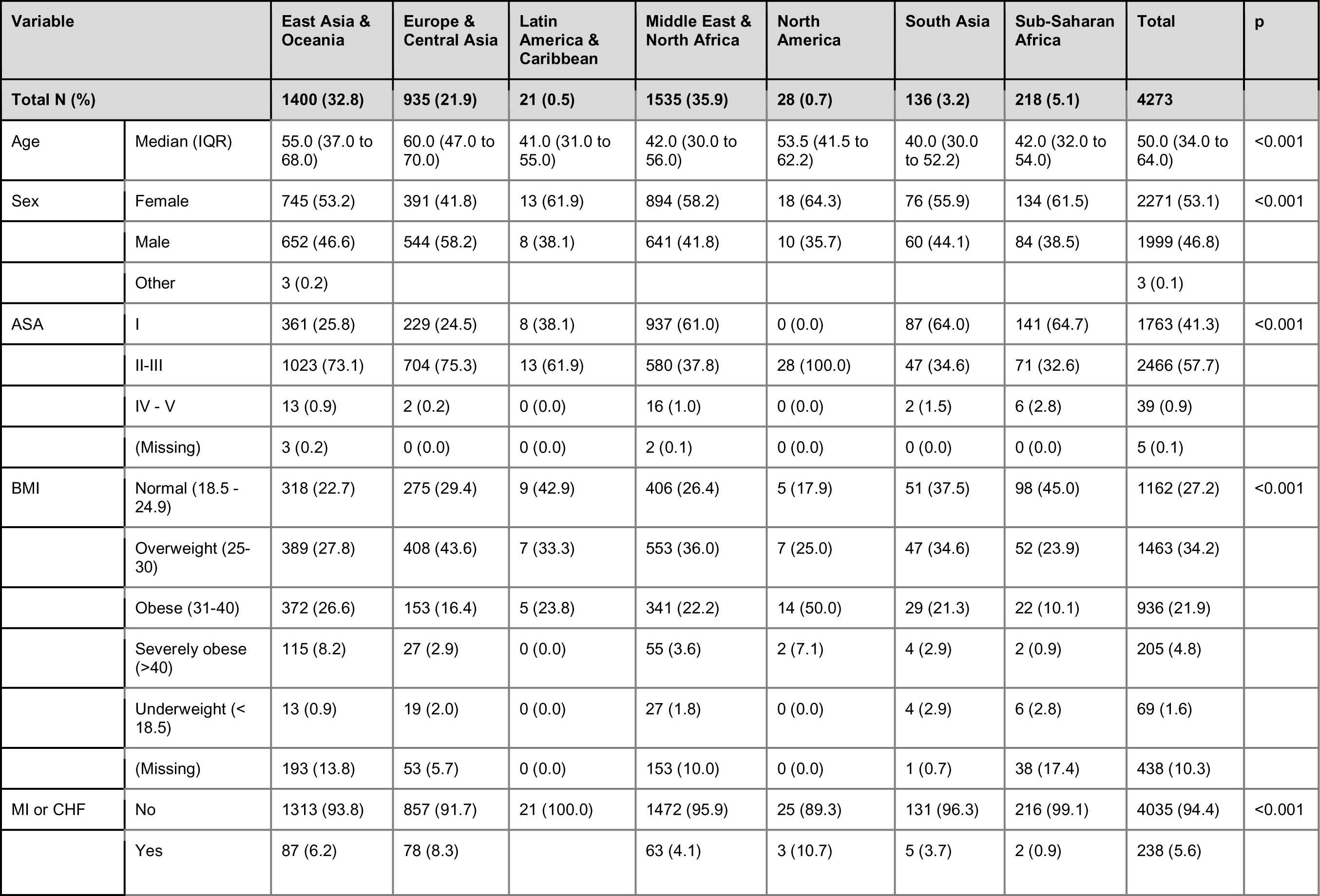
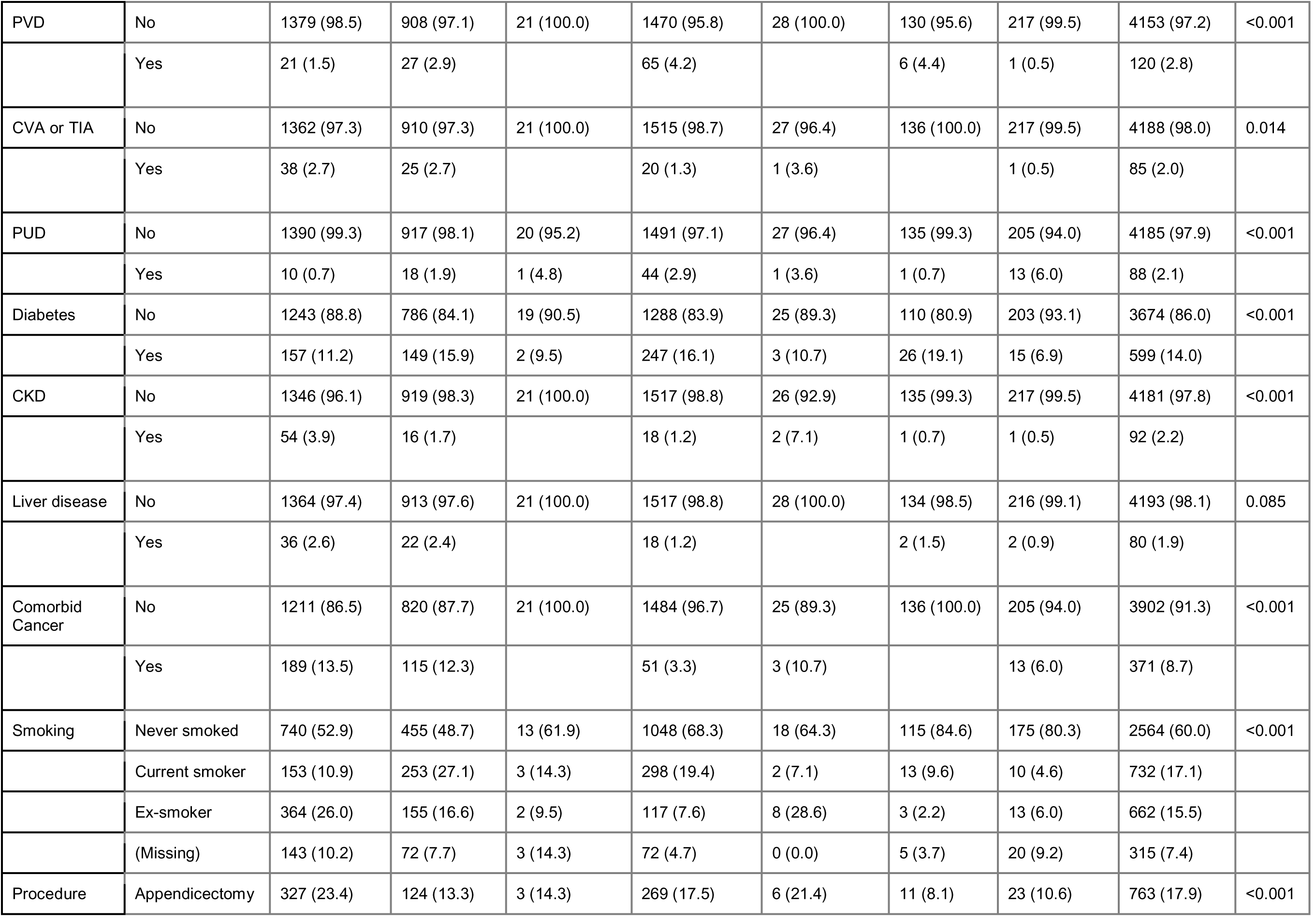
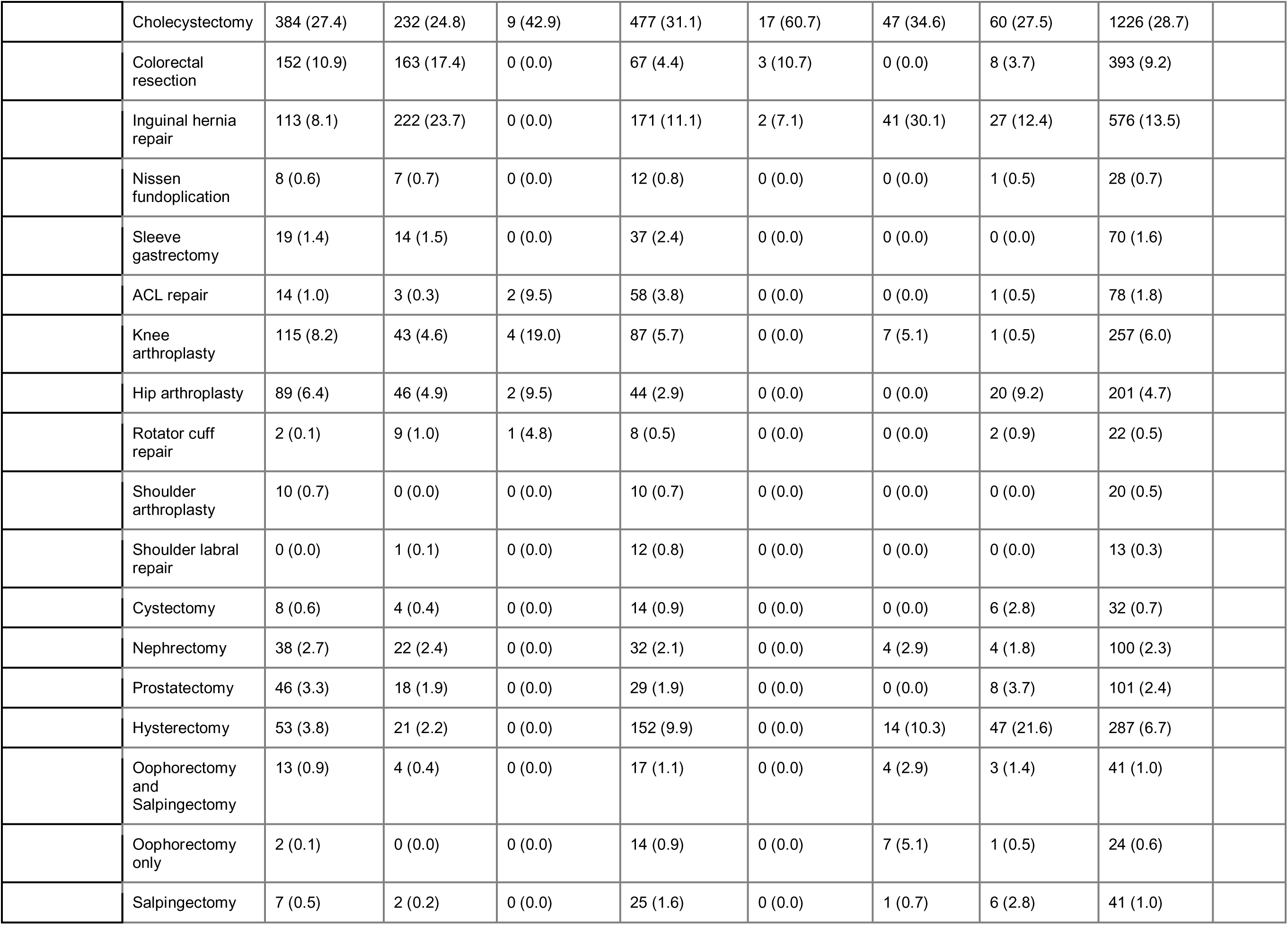
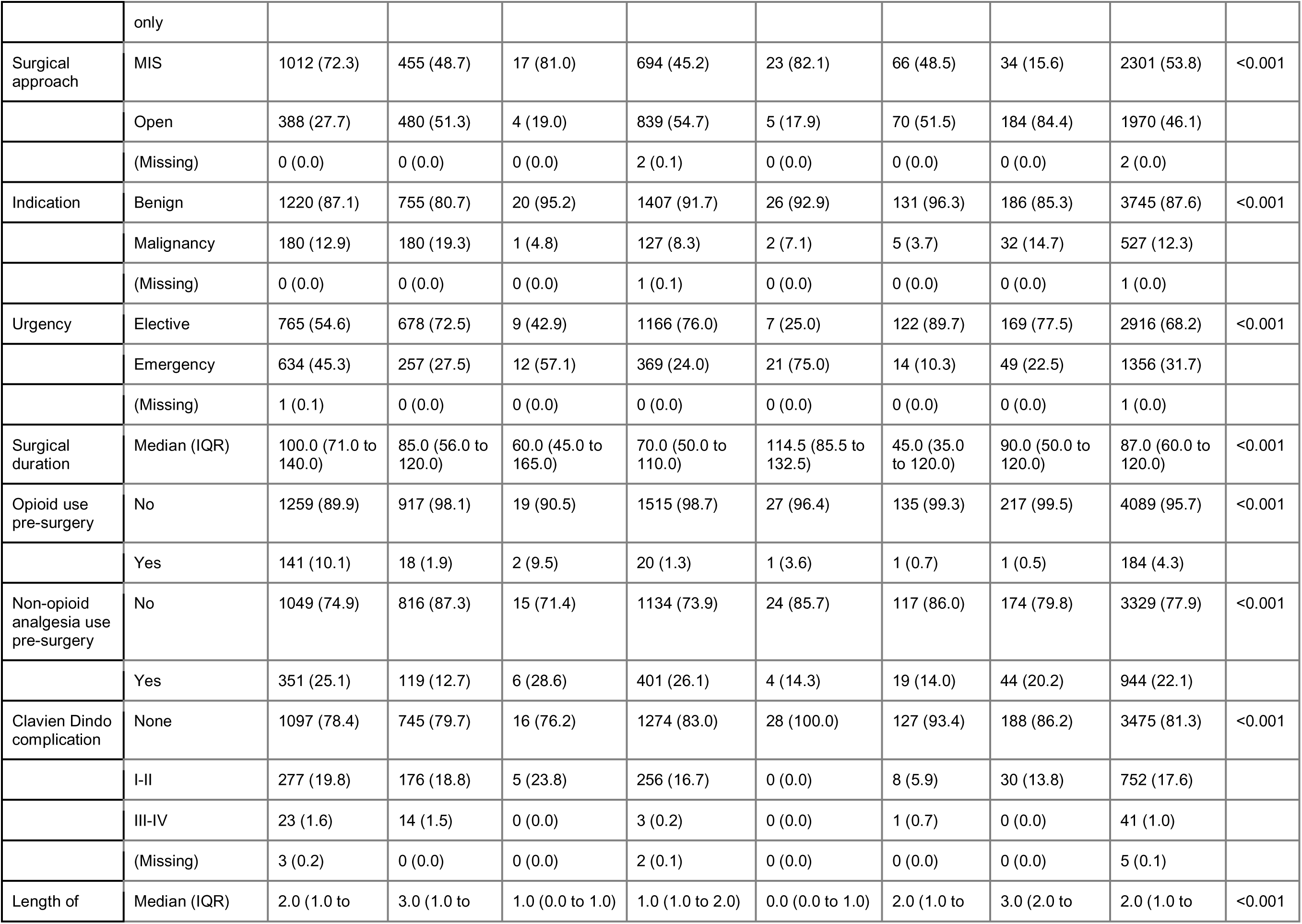
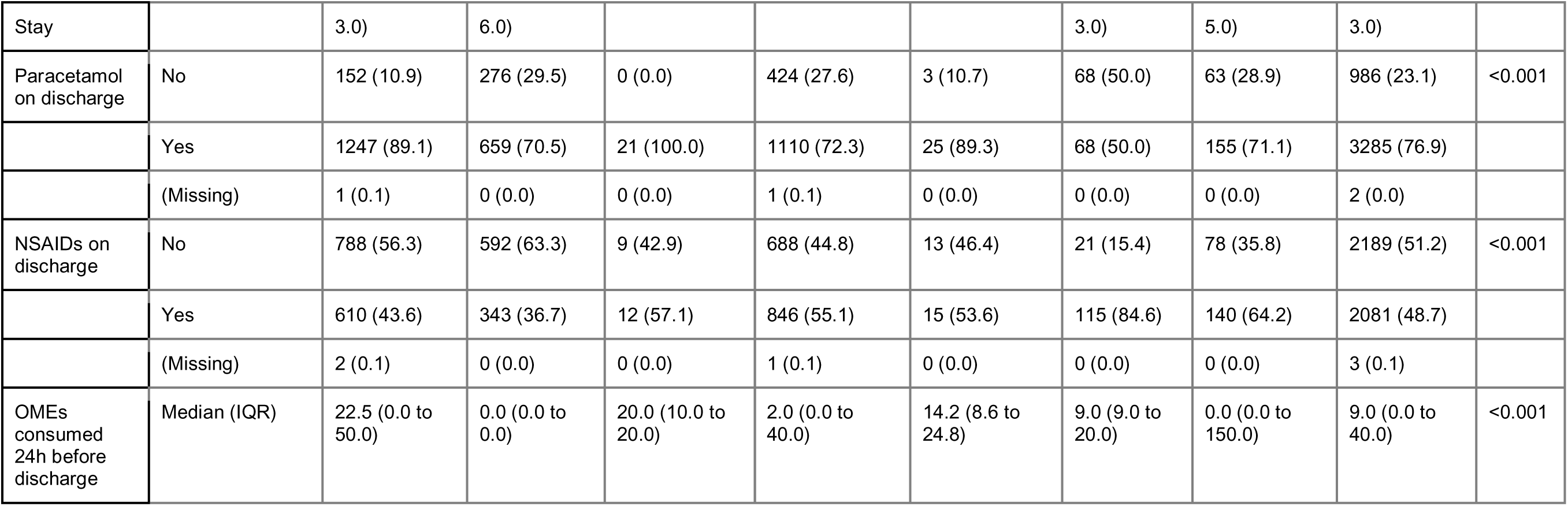
Variations in demographics by region.

**Table S12:** Patient reported outcomes by geographical region.

| Variable |  | East Asia & Pacific | Europe & Central Asia | Latin America & Caribbean | Middle East & North Africa | North America | South Asia | Sub-Saharan Africa | Total | p |
| --- | --- | --- | --- | --- | --- | --- | --- | --- | --- | --- |
| **Total N (%)** |  | **1400 (32.8)** | **935 (21.9)** | **21 (0.5)** | **1535 (35.9)** | **28 (0.7)** | **136 (3.2)** | **218 (5.1)** | **4273** |  |
| Percent time in severe pain in week post-discharge (%) | Median (IQR) | 15.0 (0.0 to 40.0) | 10.0 (0.0 to 20.0) | 10.0 (10.0 to 30.0) | 15.0 (5.0 to 35.0) | 10.0 (0.0 to 26.2) | 2.5 (0.0 to 11.2) | 11.5 (2.0 to 30.0) | 10.0 (1.0 to 30.0) | <0.001 |
| Post-discharge medical presentation for pain | No | 1197 (85.5) | 902 (96.5) | 19 (90.5) | 1344 (87.6) | 25 (89.3) | 131 (96.3) | 204 (93.6) | 3822 (89.4) | <0.001 |
|  | Yes | 203 (14.5) | 33 (3.5) | 2 (9.5) | 191 (12.4) | 3 (10.7) | 5 (3.7) | 14 (6.4) | 451 (10.6) |  |
| Post-discharge medical presentation for medication SEs | No | 1331 (95.1) | 916 (98.0) | 20 (95.2) | 1435 (93.5) | 25 (89.3) | 133 (97.8) | 211 (96.8) | 4071 (95.3) | <0.001 |
|  | Yes | 69 (4.9) | 19 (2.0) | 1 (4.8) | 100 (6.5) | 3 (10.7) | 3 (2.2) | 7 (3.2) | 202 (4.7) |  |
| Satisfaction with pain treatment (0-10) | Median (IQR) | 9.0 (8.0 to 10.0) | 9.0 (8.0 to 10.0) | 10.0 (9.0 to 10.0) | 8.0 (7.0 to 9.0) | 9.0 (8.0 to 10.0) | 9.0 (8.0 to 10.0) | 9.0 (8.0 to 10.0) | 9.0 (8.0 to 10.0) | <0.001 |
| Satisfaction amount of analgesia provided | Too little | 278 (19.9) | 119 (12.7) | 1 (4.8) | 373 (24.3) | 2 (7.1) | 29 (21.3) | 49 (22.5) | 851 (19.9) | <0.001 |
|  | Just right | 924 (66.0) | 774 (82.8) | 18 (85.7) | 1118 (72.8) | 18 (64.3) | 102 (75.0) | 159 (72.9) | 3113 (72.9) |  |
|  | Too much | 196 (14.0) | 42 (4.5) | 2 (9.5) | 41 (2.7) | 8 (28.6) | 5 (3.7) | 10 (4.6) | 304 (7.1) |  |
|  | (Missing ) | 2 (0.1) | 0 (0.0) | 0 (0.0) | 3 (0.2) | 0 (0.0) | 0 (0.0) | 0 (0.0) | 5 (0.1) |  |
| EQ-5D-5L composite score (0-100) | Median (IQR) | 80.9 (69.5 to 88.7) | 93.7 (81.3 to 100.0) | 80.9 (70.1 to 84.0) | 83.8 (73.9 to 93.7) | 83.3 (73.2 to 88.8) | 93.7 (76.1 to 100.0) | 88.7 (77.9 to 100.0) | 85.9 (73.8 to 93.7) | <0.001 |
| EQ-VAS score (0-100) | Median (IQR) | 75.0 (60.0 to 85.0) | 90.0 (75.0 to 95.0) | 90.0 (80.0 to 95.0) | 80.0 (70.0 to 90.0) | 75.0 (70.0 to 85.0) | 90.0 (80.0 to 98.0) | 90.0 (75.0 to 95.0) | 80.0 (70.0 to 90.0) | <0.001 |

## Appendix S1: Study protocol

## Appendix S2: Phone follow up script

## Appendix S3: OPERAS Opioid prescription and consumption calculations

### OPERAS Opioid Calculations

The following values were calculated for OPERAS data analysis.

A. Opioid consumption in 24 hours prior to discharge (as oral morphine equivalents (OME))
B. Total quantity of opioids prescribed on discharge (as OME)
C. Opioid consumption in 7 days after discharge (as OME)

#### A. Opioid consumption in 24 hours prior to discharge (as OME)

##### Calculation

For each opioid: Total quantity consumed (mg/mcg) x relevant conversion factor (depending on opioid type and route of administration, see **Table 1**) = OME for that opioid. Sum of all individual OMEs = total opioid consumption in 24 hours prior to discharge (as OME).

#### B. Total quantity of opioids prescribed on discharge (as OME)

##### Calculation

For each opioid: Total quantity of medication prescribed (number of tablets/ patches/ injections/ volume of liquid) x dose (mg/mcg) x relevant conversion factor = OME for that opioid. Sum of all OMEs = total opioid prescription at discharge (as OME).

#### C. Opioid consumption in 7 days after discharge (as OME)

##### Calculation

For each opioid: Total quantity of medication consumed (number of tablets/ patches/ injections/ volume of liquid) x dose (mg/mcg) x relevant conversion factor = OME for that opioid. Sum of all OMEs = total opioid consumption in 24 hours prior to discharge (as OME).

### Oral Morphine Equivalent (OME) Calculation

In order to calculate the OME for a given opioid, the opioid dose is multiplied by the relevant conversion factor (listed in Table 1).

For example, oral oxycodone can be converted to oral morphine using a conversion factor of 1.5. Therefore, the OME of oxycodone 5mg is 5 x 1.5 = 7.5mg.

OMEs with corresponding ANZCA FPM conversion factors were integrated with the REDCap data collection tool. Other OMEs were calculated using R.

Opioid combination products such as Oxycodone/ Naloxone were coded according to the opioid component.

**Table 1:** Opioid Oral Morphine Equivalent Conversion Factors used in OPERAS and corresponding source.

| Opioid | Route | Conversion factor | Source |
| --- | --- | --- | --- |
| **Morphine (mg)** | PO (tablet/ liquid) | 1 | ANZCA FPM<sup>1</sup> |
|  | SC/ IM/ IV | 3 | ANZCA FPM<sup>1</sup> |
| **Tramadol (mg)** | PO (tablet/ liquid) | 0.2 | ANZCA FPM<sup>1</sup> |
|  | SC/ IM/ IV | 0.4 | NHS<sup>4</sup> |
| **Oxycodone (mg)** | PO (tablet/ liquid) | 1.5 | ANZCA FPM<sup>1</sup> |
|  | SC/ IM/ IV | 3 | ANZCA FPM<sup>1</sup> |
| **Fentanyl (mcg)** | PO (tablet/ liquid) | 0.33 | Arnold & Weissman<sup>7</sup> |
|  | TD (patch) | 3 | ANZCA FPM<sup>1</sup> |
|  | SC/ IM/ IV | 0.2 | ANZCA FPM<sup>1</sup> |
| **Codeine (mg)** | PO (tablet/ liquid) | 0.13 | ANZCA FPM<sup>1</sup> |
|  | SC/ IM/ IV | 0.25 | ANZCA FPM<sup>1</sup> |
| **Buprenorphine (mcg)** | Sublingual | 0.04 | ANZCA FPM<sup>1</sup> |
|  | TD (patch) | 2 | ANZCA FPM<sup>1</sup> |
|  | IM/ IV | 0.075 | Nielsen et al. 2014<sup>2</sup> |
| **Pethidine (mg)** | PO (tablet/ liquid) | 0.125 | NHS<sup>4</sup> |
|  | SC/ IM/ IV | 0.4 | ANZCA FPM<sup>1</sup> |
| **Hydromorphone (mg)** | PO (tablet/ liquid) | 5 | ANZCA FPM<sup>1</sup> |
|  | SC/ IM/ IV | 15 | ANZCA FPM<sup>1</sup> |
| **Tapentadol (mg)** | PO (tablet/ liquid) | 0.3 | ANZCA FPM<sup>1</sup> |
| **Dextro-propoxyphene (mg)** | PO (tablet/ liquid) | 0.1 | ANZCA FPM<sup>1</sup> |
| **Pentazocine** | Oral | 0.16 | Stoelting's Pharmacology & Physiology<sup>6</sup> |
|  | SC/ IM/ IV | 2.5 | Stoelting's Pharmacology & Physiology<sup>6</sup> |
| **Nalbuphine** | IV | 3 | UNC<sup>5</sup> |
| **Alfentanil (mg)** | IV | 35 | NHS<sup>4</sup> |
| **Remifentanil (mg)** | IV | 33 | Arnold & Weissman<sup>7</sup> |
| **Butorphanol** | IV | 15 | UNC<sup>5</sup> |
| **Hydrocodone** | PO | 1 | UpToDate<sup>3</sup> |

#### References

1. https://www.anzca.edu.au/getattachment/6892fb13-47fc-446b-a7a2-11cdfe1c9902/PS01(PM)-(Appendix)-Opioid-Dose-Equivalence-Calculation-Table
2. https://ndarc.med.unsw.edu.au/sites/default/files/ndarc/resources/TR.329.pdf
3. https://www.uptodate.com/contents/image?imageKey=PALC%2F111216
4. https://www.gloshospitals.nhs.uk/gps/treatment-guidelines/opioid-equivalence-chart/
5. https://www.med.unc.edu/aging/wp-content/uploads/sites/753/2018/06/Analgesic-Equivalent-Chart.pdf
6. https://www.bookdepository.com/Stoeltings-Pharmacology-and-Physiology-in-Anesthetic-Practice-Pamela-Flood/9781975126896?redirected=true&utm_medium=Google&utm_campaign=Base3&utm_source=NZ&utm_content=Stoeltings-Pharmacology-and-Physiology-in-Anesthetic-Practice&selectCurrency=NZD&w=AF7CAU96V07FPHA8VTLM&gclid=EAIaIQobChMIvObWw62J-AIVzpJmAh1HVQvzEAQYBSABEgJxAfD_BwE
7. Arnold R, Weissman DE. Calculating Opioid Dose Conversions, 2nd Edition. Fast Facts and Concepts. July 2005; 36.

### PubMed Citable Authors

#### Writing group

William Xu*, Gordon Liu, Chris Varghese, Cameron Wells, Nicolas Smith, John Windsor [University of Auckland, Auckland, New Zealand], Lorane Gaborit [Australian National University, Canberra, Australia]; Sarah Goh, Aya Basam [Monash University, Melbourne, Australia]; Muhammed Elhadi [University of Tripoli, Tripoli, Libya]; Rachel Ting Qian Soh [University of Tasmania, Tasmania, Australia]; Umar Saeed [International Center of Medical Sciences Research, Islamabad, Pakistan]; Eman Abdulwahed [University of Tripoli, Tripoli, Libya]; Michael Farrell [Lehigh Valley Health Network, Allentown, United States] Deborah Wright [University of Otago, Otago, New Zealand]; Jennifer Martin, Peter Pockney*** [University of Newcastle, Newcastle, Australia]

*First author (overall guarantor)

***Senior author

#### Statistical analysis

William Xu* [University of Auckland, Auckland, New Zealand]

#### OPERAS Steering committee

Aya Basam, Sarah Goh, Jiting Li, Jainil Shah, Abdullah Waraich [Monash University, Melbourne, Australia]; Lorane Gaborit, Upasana Pathak [Australian National University, Canberra, Australia]; Amie Hilder [Deakin University, Melbourne, Australia]; Muhammed Elhadi [University of Tripoli, Tripoli, Libya]; Aiden Jabur [Griffith University, Gold Coast, Australia]; Kaviya Kalyanasundaram [University of Adelaide, Adelaide, Australia]; Christina Ohis [Western Sydney University, Sydney, Australia]; Chui Foong [Kelly] Ong [Melbourne Training Circuit, Melbourne, Australia]; Melissa Park, Venesa Siribaddana [University of Newcastle, Newcastle, Australia]; Kyle Raubenheimer [Perth Metro Training Circuit, Perth, Australia]; Jennifer Vu [Sydney University, Sydney, Australia]; Cameron Wells, Gordon Liu, Liam Ferguson, William Xu, Chris Varghese [University of Auckland, Auckland, New Zealand]

#### Scientific advisory group

Peter Pockney, Kristy Atherton, Amanda Dawson, Jennifer Martin [University of Newcastle, Newcastle, Australia]; Arnab Banerjee [Australian National University, Canberra, Australia]; Nagendra Dudi-Venkata [Royal Australasian College of Surgeons, Adelaide, Australia]; Nicholas Lightfoot [University of Auckland, Auckland, New Zealand]; Isabella Ludbrook [Hunter New England Network, Newcastle, Australia]; Luke Peters [Royal Australasian College of Surgeons, Sydney, Australia]; Rachel Sara [Counties Manukau Health, Manukau City, New Zealand]; David Watson [Flinders University, Adelaide, Australia]; Deborah Wright [University of Otago, Otago, New Zealand]

#### OPERAS National Leads

Ademola Adeyeye [Afe Babalola University Multisystem Hospital, Ado-Ekiti, Nigeria]; Luis Adrian Alvarez-Lozada [Autonomous University of Nuevo León, Monterrey, Mexico]; Semra Demirli Atici [University of Health Sciences Tepecik Training and Research Hospital, Izmir, Turkey]; Milos Buhavac [Texas Tech University Health Sciences Center, Lubbock, United States of America]; Giacomo Calini [University Hospital of Udine, Udine, Italy]; Muhammed Elhadi [University of Tripoli, Tripoli, Libya]; Orestis Ioannidis [George Papanikolaou Hospital, Thessaloniki, Greece]; Mustafa Deniz Tepe [Karadeniz Technical University, Trabzon, Turkey]; Upanmanyu Nath [Nilratan Sircar Medical College and Hospital, Kolkata, India]; Ahmad Uzair [King Edward Medical University Hospital, Lahore, Pakistan]; Wah Yang [The First Affiliated Hospital of Jinan University, Guangzhou, China]; Faseeh Zaidi, Surya Singh [University of Auckland, Auckland, New Zealand]; Bahiyah Abdullah [Hospital Universiti Teknologi MARA, Malaysia (HUiTM)], Diana Sofia Garces Palacios* [Hospital Susana Lopez De Velencia], Ahmed Ragab [Alexandria University, Alexandria, Egypt]

#### OPERAS Australian State Leads

Kyle Raubenheimer [Royal Perth Hospital, Perth, Australia]; Davina Daudu [University of Western Australia, Perth, Australia]; Sarah Goh, Simran Vinod Benyani, Nandini Karthikeyan [Monash University, Melbourne, Australia]; Laure Taher Mansour [University of Adelaide, Adelaide, Australia]; Warren Seow [University of Adelaide, Adelaide, Australia]; Zoya Tasi [University of Tasmania, Hobart, Australia]; Aiden Jabur [Griffith University, Gold Coast, Australia]; Upasana Pathak [Australian National University, Canberra, Australia]; Melissa Park [University of Newcastle, Newcastle, Australia]

#### Algeria

Dhia Errahmane Abdelmelek*, Ikram Fatima Zohra Boussahel, Oumelaz Kaabache, Naoual Lemdaoui, Oualid Nebbar [Center Anti Cancer, Sétif]; Mounira Rais*, Meriem Abdoun*, Aya Tinhinane Kouicem, Souad Bouaoud, Kamel Bouchenak, Hind Saada, Amel Ouyahia, Wassila Messai [CHU Saadna Abdennour Hospital, Sétif]

#### Australia

Zhi Shyuan Choong*, Clarissa Ting, Michelle Larkin, Pei Jun Fong, Isabel Soh, Alyssia De Grandi, Hareem Iftikhar, Akansha Sinha, Dhruv Kapoor, Tara Chlebicka [Albury Wodonga Health]; David Singer*, Kim Goddard, Lisa Matthews [Armadale Health Service, Mount Nasura]; Rosalina Lin*, Jessica Chambers, Juliet Chan, Brooke Macnab, John Barker, Morgan Mckenzie, Neil Ferguson [Armidale Rural Referral Hospital, Armidale]; Ghanisht Juwaheer*, Vijayaragavan Muralidharan, Sonia Gill, Nakjun Sung, Rohan Patel, Chris Walters, Kevin Nguyen, David Liu, Carlos Cabalag, Jennifer Lee, San-Hui Anita Leow, Suat Li Ng, Hamza Ashraf, Fraizer Mulder, Jonathan Loo, David Proud, Samantha Wong, Yida Zhou, Qi Rui Soh, David Chye, Sean Stevens, Patrick Tang, Stephen Kritharides, Jason Dong, Oscar Morice, Dora Huang, Andrew Hardidge, Mishka Amarasekara, Aleah Kink, Damien Bolton [Austin Hospital, Melbourne]; Alisha Rawal*, Jasraaj Singh*, Matthew Heard*, Yusuf Hassan*, Ahmed Naqeeb, Andrew Cobden, Duron Prinsloo, Dwain Quadros, Emma Gunn, Ha Jin Kim, Jennifer Ekwebelam, James Shanahan, Mustafa Alkazali, Mariyah Hoosenally, Naveen Nara, Peter Nguyen, Sally Barker [Ballarat Base Hospital, Ballarat]; Amie Hilder*, Ally Hui, Antara Karmakar, Bill Wang, Janindu Goonawardena, King Tung Cheung, Nicholas Chan, Ragul Natarajan, Richard Cade, Rong Jin, Shomik Sengupta, Ruth Snider [Box Hill Hospital, Melbourne]; Harsha Morisetty*, Lewis Weeda, Phoebe Sun, Lalitya Chilaka, Jacinta Cover [Bunbury Hospital]; Aashrinee De Silva Abeweera Gunasekara*, Rahavi Senthilrajan, Anas Alwahaib, Alexandra Limmer, Bushra Zamanbandhon [Campbelltown Hospital]; Kumail Jaffry [Casey Hospital, Melbourne]; Yijia Shen*, Alan Chua, Saifulla Syed [Central Gippsland Health]; Sushanth Saha*, John Glynatsis*, Lori Aitchison, Bernard Lagana, Mason Crossman, David Watson, Abby Dawson, Bryan Fong, Ella Harrison, Eleanor Horsburgh, John Glynatsis, Michael Khoo, Kritika Mishra, Lewis Hewton, Alex Mesecke, Hien Tu, Than Tun, Jason Wong [Flinders Medical Centre, Adelaide]; Elynn Ong*, Tara-Nyssa Law*, Ashlee Landy, Alyssa Leano, Andrea Li, Akshay Soni, Benjamin Dowdle, Charles Pilgrim, Dewmi Abeysirigunawardana, Deepak Rajan Jeyarajan, Diya Patel, Kyle Mckinnon, Madeline Gould, Paul Gilmore, Ruxi Geng, Rachael Loughnan, Sarahjane Norton-Smith, Solomon Nyame, Sarah Tan, Si Woo Yoon, Yantong Wang, Yichi Zhang, Zixuan Wang [Frankston Hospital]; Hans Mare*, Indrajith Withanage [Geraldton Regional Hospital]; Mitali Khattar*, Alexandra Toft, Goutham Sivasuthan, Hailin Zhao, Jordan Addley, Lucinda O’brien, Muhammad Raza, Randipsingh Bindra, Sonakshi Sharma [Gold Coast University Hospital, Southport]; Charlotte Cornwell*, Aditya Patil, Aiden Cheung, Ashleigh Lown, Amanda Dawson, Aneel Blassey, Benjamin Ochigbo, Felicity Cheng, Aleeza Fatima, Edward Zhang, Henry Kocatekin, Charles Roth, Dani Brewster, Kelvin Kwok, Paul Chen, Sharon Laura, Dominic Tynan, Edward Latif, Elizabeth Lun, Elodie Honore, Felix Ziergiebel, Jessica Blake, Karan Chandiok, Katie Bird, Lynette Ngothanh, Melissa Lee, Mariam El-Masry, Peter Hamer, Ramanathan Rm Palaniappan, Richard Mcgee, Sarah Huang, Shane Zhang, Shubhang Hariharan, Yannick De Silva, Celeste Lee, Penelope Fotheringham, Ian Incoll, Timothy Cordingley, Felicity Cheng, Matthew Brown, Leannedra Kang, Rivindu Wijayaratne, Parisse Moore, Gemma Qian, Yara Elgindy [Gosford Hospital, Gosford]; Emma Carnuccio*, Hamish Rae, Mena Shehata [Goulburn Base Hospital]; Mingchun Liu*, Brodee Lockwood, John Van Bockxmeer [Hedland Health Campus]; Ali Alsoudani*, Daniel Swan, Justin Hsieh [Ipswich Hospital]; Francesca Orchard-Hall*, Kai Yun Jodene Tay*, Raagini Mehra*, Alpha Gebeh, Ashley Bailey, Georgia Brown, Ashley Colaco, Hemashree Gopal, Jessica Boyley, Varun Changati, Joseph Fletcher, Tanishq Khandelwal, Colin House, Chris O’neil, Emily Jaarsma, Victor Ly, Zsolt Balogh, Amanda Shui, Vinogi Sathasivam, Hannah Legge-Wilkinson [John Hunter Hospital, Newcastle]; King Ho Wong*, Andrew Chen, Anthony Tran, Peter Rehfisch, Grace Wang, Jonathan Nguyen, Joshua Peker, Kayla Gallert, Mia Komesaroff, Manideep Namburi [Latrobe Regional Hospital]; Elisabeth Goldfinch*, Ropafadzo Muchabaiwa*, Aishwarya Jangam, Isobel Taylor, Iulian Nusem, Jin Hyuk (David) Park, Justin Gundara, Rachael Heigan, Tam Tran, Thomas Mackay, Yasmine Butterworth, Tomas Sadauskas, Melody Tung, Hasthika Ellepola [Logan Hospital, Meadowbrook]; Christine Gan*, Hakim Fong*, Ankita Das, Leshya Naicker, Samantha Hauptman, Aditi Kamath, Anthea Yew, Anupam Parange, Katie Kim, Sahil Kharwadkar, Tharushi Gamage [Lyell Mcewin Hospital]; Lucille Vance*, Alexandra Seldon, Moheb Ghaly [Manning Base Hospital]; Victoria Phan [Maroondah Hospital, Melbourne]; Karanjeet Chauhan*, Ahmad Bassam, Beverley Vollenhoven, Kumail Jaffry, Kajal Mandhan, Mithra Sritharan, Mahesh Sakthivel, Natalie Evans, Samuel Robinson, Seiyon Sivakumar [Monash Medical Centre, Clayton]; Liberty Marrison*, David Jollow, Krishma Joshi, Steve Tao, Pallavi Shrestha, Sai Keerthana Nukala [Northern Beaches Hospital]; Russell Hodgson*, Anna Crotty, Adriana Esho, Alasdair Harris, Amy Surkitt, Laura Bland, Blake Mcleod, Chonghao Yin, Cambo Keng, Emily Greenwood, Grace Yuan, Emma Haege, Hongyi Wu, Haotian Xiao, Isabella Pozzi, Jeff Fu, Jessica Stott Ross, Juliette Gentle, Kathy Gan, Kelvin Chang, Kexin Sun, Madhavi Singh, Maria Xie, Nicholas Mccabe, Mark Slavec, Nick Clarnette, Behzad Niknami, Peishan Zou, Sean Flintoft, Shenuka Jayatilleke, Rumnea Sok, Suqi Tan, Sanya Wadhwa, Will Swansson [Northern Hospital, Melbourne]; Daniel Abulafia*, Jian Blundell*, Amie Sweetapple, Caitlin Del Solar, Cameron Martin, David Bell, Isuru Fernando, Jared Chang, Katie Vanzuylekom, Katie Van Zuylekom, Kate Van Zuylekom, Katie Hobbs, Richard Liang [Orange Base Hospital]; Aiden Jabur*, Jazmina Tarmidi, Mahmoud Ugool, Nicholas Beatson, Sarah Bowman, Sophie Moin [Queen Elizabeth Ii Jubilee Hospital, Coopers Plains]; Wen Po Jonathan Tan*, Seevakan Chidambaram*, Siang Wei Gan, Pengnan Wang, Leshya Naicker, Katie Kim, Nicole Qiwen Wang, Yi Xin Kwan, Chinmai Patil, Divyanshu Joshi, Aditi Kamath, Aishath Hanan, Arfaan Sheriff, Jaime Duffield, Leshya Naiker, Peter Smitham, Eu Ling Neo, Matthew Chua, Shalvin Prasad, Armitesh Nagaratnam, Tarik Sammour, Yuxin Lin, Christine Lee, Eve Hopping, Muskan Jangra, Ankita Das, Ken Lin, Zachary Bunjo [Royal Adelaide Hospital, Adelaide]; Kyle Raubenheimer*, Mohamed Haseef Mohamed Yunos, Kar Long Yeung, Rachel Phu, Aisling Betts, Benjamin Just, Sahil Gera, Hilary Leeson, Jodie Jamieson, Katie Wang, Emily Luu, Michael Innes [Royal Perth Hospital]; Jennifer Vu*, Jonathan Hong, Stephen Dzator, Aki Flame, Vincent Jiang, Jianing Kwok, Aaron Lawrence, Kate Meads, Liam Pearce, Pavatharane Sarangadasa, Haylee Shaw, Victor Yu, [Royal Prince Alfred Hospital]; Elizabeth Crostella*, James Wong, Sriya Bobba, Maddison Muller, Yin Chi Hebe Hau, Thomas Wilson, Aleksandra Markovic, Jemma Green, Clara Forbes, Emalee Burrows, Lachlan Hou, Clare O’sullivan, Jonathon Foo [Sir Charles Gairdner Hospital, Perth]; Hannah Greig*, A-J Collins, Callum Chandler, Emily Heaney, Hannah Gross, Monica Morgan, Rebecca Loder, Krishnankutty Rajesh [New South Wales Local Health District Site Bega Hospital]; Shravankrishna Ananthapadmanabhan*, Akeedh Razmi, Crystal Vong, Prasanna Pothukuchi, Mary Theophilus, Roshni Sriranjan, Sharon Kaur, Marcelo Kanczuk [St John Of God Midland Public And Private Hospital, Perth]; Julia De Groot*, Angela Corrigan, Damon Li, Danniel Badri, Dominico Ciranni, Elangovan Thaya Needi, Matthew Clanfield, Nicolas Copertino, William Rumble [Sunshine Coast University Hospital]; Maria Kristina Vanguardia*, Chen Lew*, Rami Dennaoui*, Jainil Shah*, Joseph Kong, Imogen Koh, Raymond Zeng, Kristian Baziotis-Kalfas, Hannah Denby, Andy Li, Will Tran, Abhinav Singh, Olivia Lin, Michelle Chau, Olivia Donaldson, Christina (Seojung) Min, Shirahn Ballah, Sonia Ching Ting Tsui, Nathania Yong, Lucy Standish, Sarah Tan, AsukaFujihara, Lily Davies, Ramin Odisho, Anjana Ravi, Josh Collins, Pooja Chandra, Rana Abdelmeguid, GopalSingh, Xireaili Feierdaiweisi, Dharani Seneviratne, Shambhavi Srivastava, Michelle Yao, Cherilyn Teng, Nebula Chowdhury, Sasini Vidanagama, Charles Lin, Tharushi Sampatha-Waduge, Erica Wang, Chatnapa Yodkitydomying, Imogen Koh, Julia Silverii, AaronLam, Raymond Zeng, Krisha Solanki, Angus Franks, Liam Edwards, Ridvan Atilhan, Rohan Nandurkar, Oliver Wells, Kristina Vanguardia, Dennis King, Elton Edwards, Liam Edwards, Quang Tran, Michelle Chau, Seojung Min [The Alfred Hospital, Melbourne]; Abdul Rauf*, Yangzirui Fu*, Hodo Haximolla, Mengge Shang, Sharrada Segaran, Shelley Wang, Gananadha Sivakumar [The Canberra Hospital]; Jaspreet Kaur Sandhu*, Neel Mishra, Samantha Hauptman, Alyssa Chua, Danielle Chene, Guy Maddern, Henry Shaw, Qiwen Wang [The Queen Elizabeth Hospital, Adelaide]; Siyuan Pang*, Christine Lu, James Fung, Kathryn Cyr, Karen Lu, Ming Zhou How, Nelson Hu, Paul Anderson, Philip Jakanovski [The Royal Melbourne Hospital, Melbourne]; Arkan Youssef*, Howard Tang*, Rory Keenan*, Alex Chan, Mitch Canny, Farah Tahir, James Egerton, Justin Yeung, Justin Chan, Lea Tiffany, Michael Bei, Mariolyn Raj, Peter Williams, Sakshar Nagpal, Tim Outhred, Russel Krawitz [Western Health, Melbourne]; Khadijah Younus*, Mary Giurgius*, Rosemary Kirk, Amanda Gonzalez Pegorer, Pattarapan Tang-Ieam, Jack Ward, Asanka Wijetunga, Caitlin Zhang, Chris Nahm, Christine Wang, Damian Golja, Gregory Jenkins, Helena Qian, Jason Luong, Kim Nguyen, Sean Suttor, Sherman Lai, Vanessa Ma, Yan Chen [Westmead Hospital, Westmead]; Hoi Hang Yu*, Amos Lee, Antonio Barbaro, Cameron Mcguinness, Guy Maddern, Stevie Young [Whyalla Hospital & Health Services, Whyalla]; Ye Fang Lim*, Georgina Trotta, Phoebe Chao, George Ding, Carol Fang, Andi Lu, Prabhath Wagaarachchi [Women’s And Children’s Hospital]; Charlotte Cornwell*, Amy Gojnich, Peter Stewart, Isabella Dong, Kenneth Wong, Luca Burruso, Lucinda Hogan, Nathan Mcorist, Ramnik Singh, Ragavi Jeyamohan, Zhen Hou, William Lai, Emily Taylor [Wyong Public Hospital, Wyong]

#### Colombia

Diana Sofia Garces Palacios*, Maria Alejandra Nanez Pantoja, Daniel Mauricio Bolanos Nanez, Gilmer Omar Perez Hernandez, Lia Jasmin Jimenez Ramirez [Hospital Susana Lopez De Velencia]

#### Egypt

Mohamed Mohamed*, Ahmed Kamal El-Taher, Ahmed Elewa, Mahmoud Ayman Soliman, Menna Diab, Radwa Ali [Al Tayseer Hospital, Zagazig]; Ahmed Ahmed*, Adham Galal, Ahmed Elkhodary, Ali Alaa, Arwa Faisal, Asmaa Badawy, Donia Eldomiaty, Mohamed Al Sayed, Esraa Rasslan, Mohamed Ramadan, Gamal Elsayed Fares, Hashem Altabbaa, Humam Emad, Muneera Alboridy, Mahmoud Mongy, Osama Albarhomy, Osama Selim, Rawan Rafaei, Raneem Atta, Ahmad Altaweel, Yara Sherif, Youssef Elghoul, Yousef Tarek [Alexandria Main University Hospital, Alexandria]; Dina Atef*, Ahmed Mahmoud*, Mahmoud Saad*, Mohamed Ragab, Aya Hussien, Mostafa Abdelbaky, Ismail Muhammad, Afnan Morad, Ahmed Ali, Ahmed Hussien, Ahmed Shipa, Ahmed Aboulfotouh, Ahmed Mohamed Hashem, Ahmed Morsi, Alshymaa Ebrahim, Ahmed Mohamed Sayed, Amira Abdelrahman, Aml Ali, Samah Abdelnaeam, Asmaa Emam, Aya Shaban, Fady Barsoum, Esraa Mostafa, Doaa Abdelbaset, Dina Othman, Safaa Othman, Nour Salah Khairallah, Salma Morsi, Armia Azer, Enas Abdelbaset Abdelsamed, Islam Ibrahim, Esraa Abdelbaset, Esraa Hamoda, Fatma Monib, Fatma Harb, Hager Maher, Haitham Mohammed, Kerollos Henes, Kerollos Shamshoon, Mahmoud Hassanein, Magdy Mahdy, Mahmoud Khalil, Manal Ali, Mansour Khalifa, Marwa Amary, Merna Ezz Suliman, Mohammed Saif Al Nasr, Michael Elia, Michael Adly, Mo’men Roshdy, Mohammed Al-Quossi, Mohammed Fargaly, Mona Saber, Mostafa Abbas, Ola Haroon, Omima Khalil, Omnia Talaat, Rahma Elnagar, Randa Soliman, Reham Aboelela, Salem Salah, Samia Abdelgawad, Tarek Hussien, George Sobhy, Yasmeen Sayed, Yousra Othman [Assiut University Hospital, Assiut]; Reham Silem*, Ali Dawood, Tarek Hemaida, Reem Ahmed [Aswan University Hospital, Aswan]; Ebrahim Salem, Osama Fathy Ali Ali Rashed, Mohamed Halawa [El Tadamon Specialised Hospital, Portsaid]; Hossam Elfeki*, Abdelrahman Mosaad, Abdelrahman Shaaban, Hebatalla Abdelsalam, Ahmed Sakr, Aly Sanad, Amr Elsawy, Bassant Maged Maged, Dana Hegazy, Mohamed Abdelmaksoud, Mahmoud Laymon, Mohamed Taman, Esraa R Moawad, Hadeer Aboelfarh, Karim Elkenawi, Manar Osama, Mirna Sadek, Mohamed Abdelaziz Elghazy, Mohammed Attiah, Mohamed Nader, Mostafa Shalaby, Omar Attiya, Osama Samir Gaarour, Ahmed Zaghloul [Mansoura University Hospital, Mansoura]; Pola Mikhail*, Karim Badr, Hatem Soltan, Mohamed Donia, Mohammed Gaafar [Menofia University Hospital, Menofia]; Khaled Abdelwahab*, Abdelaziz Sallam, Ahmed Eid, Mohamed Yousri, Omar Hamdy [Oncology Center Mansoura University, Mansoura]; Aiman Al-Touny*, Abdelrhman Alshawadfy, Ahmed Hamdy, Ahmed Ellilly, Ahmed Mahdy, Ahmed El-Sakka, Hamdy Hendawy, Asmaa Salah, Bassma Raslan, Eman Teema, Eslam Albayadi, Esraa Nasser, Hanaa Mohamed, Mohamed Mahmoud, Mostafa Elsaied, Omima Taha, Shaimaa Dahshan, Shimaa Al-Touny, Ahmed Karrar, Ahmed Khairy [Suez Canal University Hospital, Ismailia]; Alaa Mohamed Ads*, Rabiaa Alomar, Issa AbuShawareb, Abdallah Saeed, Abdelhafeez Mashaal, Adel Mohamed Ads, Sohila Ghanem, Ahmed Elghamry, Eman Ayman Nada, Youssef Ali Noureldin, Mohamed Fayez Fouda, Nourhan Shaheen, Shereen Allam, Ibrahim Mazrou, Ali Fahmy Shehab, Wesam Kussaili [Tanta University Hospital, Tanta]

#### Greece

Dimitrios Korkolis*, Evangelos Fradelos, Aikaterini Sarafi [Agios Savvas Anticancer Hospital Of Athens]; Nikolaos Machairas*, Konstantinos S. Giannakopoulos, Fotios Stavratis, Georgios Korovesis, Gerasimos Tsourouflis, Myrto D. Keramida, Nikolaos Kydonakis, Stylianos Kykalos, Athanasios Syllaios, Panagiotis Dorovinis, Dimitrios Schizas [General Hospital Of Athens “Laiko”]; Orestis Ioannidis*, Anastasia Malliora, Elissavet Anestiadou, Konstantinos Zapsalis, Fotios Kontidis, Lydia Loutzidou, Nikolaos Ouzounidis, Stefanos Bitsianis, Savvas Symeonidis, Smaragda Skalidou, Orestis Ioannidis, Olga Maria Valaroutsou [General Hospital Of Thessaloniki “George Papanikolaou”]; Themistoklis Dagklis*, Alexandra Arvanitaki, Apostolos Mamopoulos, Apostolos Athanasiadis, Stergios Kopatsaris, Ioannis Kalogiannidis, Ioannis Tsakiridis, Georgios Kapetanios, Evangelos Papanikolaou, Nikolaos Tsakiridis, Fotios Zachomitros [Hippokratio General Hospital Of Thessaloniki]; Andreas Larentzakis*, Argyrios Gyftopoulos, Konstantinos Albanopoulos, Apostolos Champipis, Christos Yiannakopoulos, Gavriella Zoi Vrakopoulou, Konstantinos Saliaris, Konstantinos Lathouras, Spyridon Skoufias, Georgia Doulami [Iaso]; Metaxia Bareka*, Eleni Arnaoutoglou, Fragkiskos Angelis, Fragkiskos Angeslis, Michael Hantes, Maria Ntalouka [Larissa University Hospital]

#### Iraq

Maytham A. Al-Juaifari*, Mohammed Alwash, Rasool Maala, Yasir Adnan Zwain, Sara Ahmed Saleh, Mohammed Khorsheed [Al-Najaf Al-Ashraf Teaching Hospital, Najaf]

#### Italy

Antonio Pesce, Carlo V. Feo*, Massimiliano Bernabei*, Francesca Petrarulo, Nicolò Fabbri, Raffaele Labriola, Silvia Jasmine Barbara [Azienda Unità Sanitaria Locale di Ferrara-University of Ferrara, Ferrara]; Simone Bosi*, Angela Romano, Anna Canavese, Caterina Catalioto, Claudio Isopi, Cristina Larotonda, Gerti Dajti, Matteo Rottoli, Iris Shari Russo, Stefano Cardelli [Irccs Azienda Ospedaliero, Bologna]; Francesco Castagnini*, Francesco Traina, Giulia Guizzardi, Giulia Giuzzardi, Mara Gorgone, Marco Maestri [IRCCS Istituto Ortopedico Rizzoli, Bologna]; Pasquale Cianci*, Ivana Conversano, Enrico Restini, Domenico Gattulli, Giorgia Grillea, Marco Varesano [Lorenzo Bonomo, Andria]; Giacomo Calini*, Adelaide Andriani, Davide Gattesco, Giovanni Terrosu, Mattia Zambon, Pietro Matucci Cerinic, Luisa Moretti, Davide Muschitiello, Samantha Polo, Vittorio Bresadola [University Hospital Of Udine, Udine]

#### Jordan

Salah Abu Wardeh**\***, Mahmoud Al-Baw*, Saif Alhaleeq*, Subhi Al-Issawi*, Abdalqader Al Smadi, Esmat Alsaify, Farah Banihani, Noor Massadeh, Nada Massadeh, Dima Al-issawi, Basel Elyan, Qotadah Al-Shami, Yazan Alomari [Al Basheer Hospital, Amman]; Abed Alazeez Alkhatib*, Bader Alzghoul, Ahmad Saleh, Jamal Yaghmour, Mahmoud Shahin, Mohammed Maali [Al Istiklal Hospital, Amman]; Dawood Alatefi*, Heba Al-Smirat, Abdulhakim Hezam, Nassar Alathameen [Alkarak Governmental Hospital, Alkarak]; Amr Al Hammoud*,Abdulrahim Al Kaddah*, Salem Ayasrah, Hamza Abuuqteish, Tesneem Al-Mwajeh, Reena Makableh, Saad Bataineh, Amin Shabaneh, Wesam Alnatsheh, Marwan Aldeges, Huda Hamad, Sireen Shehahda, Dima Khassawneh, Osama Alzyoud, Risan Alrosan, Hasan Awad, Tariq Khaldoon, Rabab Shannaq, Mohammad Al hamoud, Bader Abo fadalah, Mo’ath Al-Hazaimeh, Wail Khraise [King Abdullah University Hospital, Irbid]; Lara Alnajjar*, Majjd Alnajjar*, Sohaib Al-Omary*, Adnan Ababneh, Alaa Albashaireh, Mohammad Khadrawi, Mohammad Aljamal*, Tayseer Athamneh, Ro-a Muqbel, Maryam Al-jammal, Ahmad Masarrat, Alia Al-zawaydeh, Ibrahim Taha, Taima’ Qattawi, Rayyan Smadi, Ayah Alhaleem, Mosab Alboon, Omar Hazaymeh, Leen Karasneh, Safa’ Al-Haek [Princess Basma Teaching Hospital, Irbid]

#### Libya

Marin Almahroush*, Tamam Alfrijat, Aya Elporgay, Hadeel Shanag, Hamza Agilla, Hind Alameen, Marya Bensalem, Mawadda Altair, Malak Ghemmied, Rehab Alarabi, Sara Alhudhairy [Abu Saleem Trauma Hospital, Tripoli]; Rima Gweder*, Amal Alzarroug, Ebtihal Alabed, Fadwa Elreaid, Omar A Elkharaz, Fatma Fathi Elreaid, Safa Sasi Albatni [Alkhadra Hospital, Tripoli]; Haitham Elmehdawi*, Milad Gahwagi*, Ayman Mohamed, Tariq Alfrjani, Khaled Khafifi, Ayat Rasheed, Ayoub Akwaisah, Hassan Bushaala, Mustafa Elfadli, Mohamed Moftah, Salima Algabbasi, Salma Esaiti, Sara Elfallah, Abtisam Alharam, Fatima Alariby, Mohamed Isweesi, Tarik Ahmed Eldarat, Ayman Arhuma Dabas [Benghazi Medical Center, Benghazi]; Akram Alkaseek*, Ahmed Mohammed Abodina, Aya Alqaarh, Hibah Bileid Bakeer, Hoda Salem Alhaddad, Husein Aboudlal, Sawsan Alsaih [Gharyan Central Hospital, Gharyan]; Noora Abubaker, Najwa Abdelrahim*, Ali Alzarga, Basma Omar, Farah Faris, Qamrah Alhadad [Ibn Sina Teaching Hospital, Sirt]; Asma Abufanas*, Hussameddin Badi*, Israa Benismai*, Hawa Obeid*, Abdulwahab Abdalei, Ahmed Abdulrahman, Aisha Swalem, Ebtisam Alzarouq, Amna Safar, Esra Shagroun, Boshra Hashem, Fatheia Elrishi, Fatima Abdulali, Habeeba Ahmed, Ibrahim Eltaib, Joma Elzoubia, Aisha Albarki, Hoda El Mugassabi, Fatima Abushaala, Amany Abuzaho, Nida Juha, Raneem Egzait, Sundes Shetwan, Alzahra Lemhaishi, Faisel Matoug [Misurata Central Hospital, Misurata]; Eman Abdulwahed*, Aamal Askar, Abir Ben Ashur, Adel Bezweek, Bushra Altughar, David Emhimmed, Donia Elferis, Laila Elgherwi, Enas Soula, Doaa Gidiem, Maren Grada, Khawla Derwish, Maram Alameen, Nassib Algatanesh, Ahlam Elkheshebi, Reem Ghmagh, Sharf Barka, Sultan Ahmeed, Sarah Aljamal, Zahra Alragig, Mohamed Addalla, Ahmed Atia, Atab Kharim, Fathia Mahmoud, Muhannud Binnawara, Entisar Alshareea [Tripoli Central Hospital, Tripoli]; Mohamed Alsori*, Aisha Alshawesh, Ghaliya Mohamed H Alrifae, Amira Ashour, Anwaar Abozid, Asil Omar Saleh Alflite, Anwar Mohamed, Jaber Arebi, Fatma Alagelli, Hana Yousef Gineeb, Rawia Ghmagh, Rihab Mohammed Bin Omar, Retaj Alaqoubi, Sara mohammed, Serien Hossain Bensalem, Tahani Elgadi, Wesam Sami, Yara Bariun, Abdulhadi Mohammed Alhadi Alhashimi, Dheba Almukhtar Abdulla, Heba Rhuma, Husam Enaami, Asraa Ali Alboueishi [Tripoli Medical Center/ Tripoli University Hospital, Tripoli]; Hayat Ben Hasan* [Zliten Medical Centre, Zliten]

#### Lithuania

Narimantas Samalavicius*, Vitalijus Eismontas, Jonas Jurgaitis, Oleg Aliosin, Vitalija Nutautiene [Klaipeda University Hospital]

#### Malaysia

Andee Dzulkarnaen Zakaria*, Anil Kumar Sree Kumar Pillai, Dinesh Kumar Vadioaloo, Mohamed Ashraf Mohamed Daud, Jien Yen Soh, Mohd Zaim Zakaria [School of Medical Sciences & Hospital USM, Universiti Sains Malaysia]; Siti Mayuha Rusli*, Nur Ayuni Khirul Ashar*, Zatul Akmar Ahmad*, Afiq Aizat Ramlee, Sharifah Nor Amirah Syed Abdul Latiff Alsagoff, Ahmad Anuar Sofian, Muhammad Badrul Hisyam Mohamad Jamil, Bahiyah Abdullah, Mohamad Faiz Noorman, Muhammad Fihmi Zainal Abidin, Mohamed Izzad Isahak, Siti Nasyirah Nisya Adnan, Zaidatul Husna Mohamad Noor, [Hospital Universiti Teknologi Mara (HUiTM)]

#### Mexico

Luis Adrian Alvarez-Lozada*, Alejandro Quiroga Garza, Andrea Aguilar Leal, Bernardo Alfonso Fernández Reyes, Ethel Valeria Orta Guerra, Francisco Javier Arrambide Garza, Héctor Erasmo Alcocer Mey, Jorge Arath Rosales Isais, Juventino Tadeo Guerrero Zertuche, Patricia Ludivina González García, Luis Antonio Heredia Sánchez, Marcela Patricia Flores Mercado, Oscar Alonso Verduzco Sierra, Pedro Emiliano Ramos Morales, Stephie Oyervides Fuentes, Víctor Manuel Peña Martínez [University Hospital Dr. Jose Eleuterio Gonzalez, Monterrey, Nuevo Leon]

#### New Zealand

Surya Singh*, Arwa Hadi, Christian Woodbridge, David Thornton-Hume, Jack Forsythe, Isini Dharmaratne, Vivian Pai, John Windsor, Kamran Zargar, Lucy Waldin, Lily Winthrop, Matias Alvarez, Meileen Huang, Matt Kumove, Marta Simonetti, Namisha Chand, Oliver Goldsmith, Oscar Guo, Paul Monk, Karen Zhou, Sai Harshitha Penneru, Shaamnil Prasad, Seifei Ren, Terrence Hill, Vyoma Mistry, Selena Sun [Nz, Auckland, Auckland City Hospital]; Ashley Pereira*, Scott Mclaughlin*, Andrew Stokes, Avinash Sathiyaseelan, Jeremy Rossaak, Janice Lim, Kenya Brooke, Liam Quinlan, Mark Pottier, Nayanika Podder, Puja Jinu, Shanay Ramphal, Wikus Vermeulen [Nz, Bay Of Plenty, Tauranga Hospital]; Fraser Jeffery, Ibrahim S. Al Busaidi, Janelle Divinagracia, William Ju, Yizhuo Liu, Tamara Glyn, Nasya Thompson* [Nz, Canterbury, Christchurch Hospital]; Vivien Graziadei*, Joshua Canton*, Joseph Furey*, Horim Choi, Grace Coomber, Tanya Divekar, Tessa English, Erin Gernhoefer, Tom Healy, Justin Chou, Dikshya Parajuli, Catherine Reed, Rod Studd, Anthony Lin [Nz, Capital And Coast, Wellington Hospital]; Cameron Wells*, Cindy Xu*, Arwa Hadi, Andrew Maccormick, Heejun Park, Athulya Rathnayake, Brittany Williams, Ashley Chan, Corinne Smith, Francesca Casciola, Jainey Bhikha, Jonathan Luo, Kevin Yi, Megan Singhal, Ria George, Rosie Luo, Taylor Frost [Nz, Counties Manukau, Middlemore]; Fatima Hakak*, Akhita George, Angela Carlos, Annie Ho, Connor Mcrae, Jonathan Lescheid, Jenny Soek, Andrew Pham, Sophie St Clair, Su-Ann Yee, Jennifer Lim [Nz, Lakes, Rotorua Hospital]; Taehoon Kim*, Anne Qi Chua, Christopher Harmston, Hamish Boyes, Holly Cook, Jamie Struthers, Jess Radovanovich, Nicholas Quek [Nz, Northland, Whangarei Base Hospital]; Chekodi Fearnley-Fitzgerald*, Deborah Wright, Kushan Ghandi, Natalie Matheson [Nz, Southern, Dunedin Hospital]; Matthew James McGuinness*, Brian Chen, Rebecca Indiana Douglas, Konrad Richter, Nisha Bianca Soliman, Scott Matthew Bolam, Vineeth Vimalan, William Currie [Nz, Southern, Invercargill (Kew) Hospital]; Mitchell Cuthbert*, Poppy Ross*, Amy Nicholson, Briar Garton, Emilie Agnew, Niamh Conlon, Nicholas Waaka, Ritwik Kejriwal, Sean Nguyen, Edmund Leung [Nz, Taranaki, New Plymouth Hospital]; Milidu Ratnayake*, Quintin Smith*, Nejo Joseph*, Bosco Yue, Calvin Fraser, Charles Lam, Ethan Figgitt, Gordon Liu, Kevin Tan, Ha Seong You, Helen Zheng, Jenny Luo, James Sharp, Kabir Khanna, Levi Simiona, Michel Luo, Milidu Ratnayake, Patrick Wong, Rebecca Luu, Rohit Paul, Shiva Nair, Shadie Asadyari-Lupo, Wing Hung, Geoffrey Ying [Nz, Waikato, Waikato Hospital]; Jess Ho*, Alan Wu, Eamon Walsh, Jouyee Lee, Jessie Liu, Sunny Yao, Omar Nosseir, Jennifer Dang, Simon Young, Sof’ya Zyul’korneeva, Theresa Boyd [Nz, Waitemata, North Shore Hospital]; Jess Ho*, Alan Wu, Sunny Yao, [Nz, Waitemata, Waitakere Hospital]

#### Nigeria

Abdullahi Musa Kirfi*, Adamu Bala Ningi, Mohammad Albuhari Garba, Makama Baje Salihu, Ohia Ernest Ukwuoma, Abdullahi Ibrahim, Isa Mienda Sajo, Muhammad Baffah Aminu, Liman Haruna Usman, Oloko Nasirudeen Lanre, Ibrahim Shaphat Shuaibu, Stephen Yusuf, Tiamiyu Ismail, Gabi Ibrahim Umar [Abubakar Tafawa Balewa University Teaching Hospital Bauchi, Bauchi]; Ademola Adeyeye*, Ehis Afeikhena, Favour Nnaji, Joy Agu, Temi Maxwell, Tosin Motajo, Karo Ifoto, Seubong Okon [Afe Babalola University Multisystem Hospital, Ado Ekiti]; Jerry Godfrey Makama*, Amina Abosede Mohammed-Durosinlorun, Bashiru Aminu, Polite Iwedike Onwuhafua, Caleb Mohammed, Lubabatu Abdulrasheed, Joel Amwe Adze, Khadijah Richifa Suleiman, Lydia Regina Airede, Mathew Chum Taingson, Stephen Bodam Bature, Stephen Akau Kache, Uchechukwu Ohijie Ogbonna [Barau Dikko Teaching Hospital, Kaduna]; Mohammed Bello Fufore*, Abdulkarim Iya, Adeshina A Ajulo, Ahmad Mahmud, Bilal Shuaibu Yahya, Farida Onimisi-Yusuf, Hope Isaac, Timothy Jawa, Fashe Joseph, Bemi Kala, Maisaratu A Bakari, David Wujika Ngwan, Abubakar Umar, Abraham L Filikus, Daniel Wycliff [Modibbo Adama University Teaching Hospital, Yola]; Abiodun Okunlola*, Olukayode Abiola, Adebayo Adeniyi, Olabisi Adeyemo, Babatunde Awoyinka, Olakunle Babalola, Adewumi Bakare, Taiwo Buari, Cecilia Okunlola, Gbadebo Adeleye, Adedayo Salawu, Henry Abiyere, Adetolu Ogidi, Tesleem Orewole [Federal Teaching Hospital, Ido Ekiti]; Habiba Ibrahim Abdullahi*, Godwin Akaba, Arome Achem, Asi-oqua Bassey, Emeka Ayogu, Bilal Sulaiman, Dennis Anthony Isah, Chukwunonso Nnamdi Akpamgbo, Felicia Asudo, Nathaniel Adewole, Omachoko Oguche, Peter Ejembi, Samuel Ali Sani, Paul Chimezie Andrew, AliyuYabagi Isah, Bolarinwa Eniola, Zumnan Songden, Teddy Agida, Terkaa Atim [University Of Abuja Teaching Hospital, Gwagwalada]; Taofiq Olayinka Mohammed*, Hadijat Olaide Raji*, Femi Ibiyemi, Hafeez Salawu, Olushola Fasiku, Remi Sanyaolu Solagbade, Mariam Motunrayo Shiru, Gbadebo Hakeem Ibraheem, Justina Oruade, Grace Ezeoke [University Of Ilorin Teaching Hospital, Ilorin]

#### Pakistan

Tabish Chawla*, Aliya Begum Aziz, Anoosha Marium, Ayesha Akbar Waheed, Faiqa Binte Aamir, Faiza Qureshi, M Hammad Ather, Iqra Fatima Munawar Ali, Izza Tahir, Maha Ghulam Akbar, Ronika Devi Ukrani, Sajjan Raja, Sehar Salim Virani, Shahryar Noordin, Saif Ur Rehman, Shalni Golani, Syed Roohan Aamir, Syed Musa Mufarrih, Usama Waqar, Maliha Taufiq [Aga Khan University Hospital]; Ahmed Siddique Ammar*, Adya Ejaz*, Albash Sarwar, Ahmed Usman Khalid, Shehrbano Khattak [Bahria International Hospital Lahore]; Aliza Imran, Omer Bin Khalid, Urauba Kaleem, Urwah Muneer, Yumna Kashaf [Creek General Hospital]; Fatima Zafar*, Adil Zaheer, Muhammad Ali, Amna Shafaat, Arisha Qazi, Asjad Imran,Mahnoor Tariq, Muhammad Nadeem Aslam, Shehroz Ali, Tabish Atiq, Tayyiba Wasim, Daniyal Babar, Ahmad Zain, Muhammad Ibtisam [Services Hospital Lahore]; Uzair Ahmed, Syed Talha Bin Aqeel, Muhammad Muhib, Muhammad Anas Abbal, Nasar Ahmad Khan, Imran Javed [United Hospital]

#### Palestine

Layth Alkaraja*, Dana Amro, Ghaida Manasrah, Ibraheem Hammouri, Ihab Abu Hilail, Jihad Zalloum, Laith Alamlih, Mahmoud Nasereddin, Munia Rajabi, Sa’ed Shalalfeh, Zeinab Natsheh [Hebron Government Hospital, Hebron]; Khamis Elessi*, Mustafa Abu Jayyab*, Mohammed Astal, Mosheer Al-Dahdouh [Nasser Medical Complex, Gaza]; Alaa Eddin Salameh*, Alaa Ayyad, Nimatee Dawod, Hamza Alsaid, Iyas Matar, Majd Hassan, Mohammed Bakeer, Mohammad Malasah, Shehab Abuhashem, Mohammed Salem, [Palestine Medical Complex, Ramallah]

#### Romania

Sorinel Lunca*, Mihail Gabriel Dimofte, Stefan Morarasu, Ana Maria Musina, Cristian Ene Roata, Natalia Velenciuc [Regional Institute Of Oncology Iasi, Iasi]

#### Russia

Aleksandr Butyrskii*, Maxim Bozhko, Amet Ametov [Emergency Municipal Hospital No.6]

#### Saudi Arabia

Sharfuddin Chowdhury*, Doaa Bagazi [King Saud Medical City, Riyadh]

#### Spain

Julio Domenech*, Alejandro Rosello-Añon, Ana Monis, Caterina Chiappe, Beatriz Cuneo, Pablo Clemente-Navarro, Jorge Febre, Jorge Sanz-Romera, Marcos Lopez-Vega, Ignacio Miranda, Rocio Valverde-Vazquez, Sara Garcia, Maria Jose Sanguesa, Zutoia Balciscueta [Hospital Arnau De Vilanova]; Enrique Ruiz*, Eduardo Marco, Elena Talavera, Joan Farre, Loreto Bacariza, Mireia Duart, Violeta Ureña, Xenia Carre [Hospital Sant Joan Reus]

#### Sudan

Hytham K. S. Hamid*, Montasir A. Abd-Albain, Sami Galal-Eldin [Al-Moalem Medical City]; Monira Sarih*, Eithar Adam, Samir Ismail, Malaz Azhari, Tawfieg Hassan [Alandalus Clinic, Elduiem]; Mohamed Salaheldein*, Zainab Abdalla, Wahiba Ahmed [Bashair Teaching Hospital, Khartoum]; Monzer Abdulatif Mohamed Alhassan*, Hozifa Mohamed Abdalla Suliman, Hozifa Mohamed Bdalla Suliman [Education Karima Hospital, Merowe]; Rogia Ahmed Abdalla Ahmed*, Enas Mohammedtom Abdulhameed Babekir, Munya Ali Talab Khairy, Maha Mukhtar Ahmed Mukhtar, Rzan Ali Hamedelneel Ali, Yasir Babkir Ali Al-Shambaty [Elduiem Teaching Hospital, Elduiem]; Fatima Imad Yousif*, Hawa Mohammed Hassan Mohammed, Lana Osher, Lana Osher, Menhag Abdelbast, Mohamed Yassin, Noon Moawia, Rowa Abdalsadeg [Gadarif Teaching Hospital, Gadarif City]; Abrar Husein, Baraa Elhassan, Alnazeer Y. Abdelbagi, Mohammed A. Adam, Eithar M. Ali, Ibrahim A.b. Mohammed, Maab Mohamed, Mohamed Abdulaziz, Mazin Akasha, Muaz Hassan, Nadir Hilal, Noon Abdalla Abdelrahman Mohamed, Noora Abubaker, Omeralfarouk Mohammed, Shakir Mohamed, Walaa Osman, Fatima Mustafa, Alaa A Salih [Ibn-Sina Hospital, Khartoum]; Doua Ali*, Doha Mohammed Ahmed Almakki, Hanan Elnour Mohamed, Abdelhadi Elmubark, Mohamed Hassan, Ammar Alnour, Amna Elaagib, Ayman Abdelrahman, Mubarak Abdelkhalig, Khalid Nour Eldaim, Afra Babiker, Entisar Ahmed, Maab Ali, Eman Hussain, Mansour Wedatalla, Alaaaldeen Ahmed, Alla Aldeen Hamza, Mohab Mohammed, Omer Osman, Reham Ibrahim, Rihab Ahmed, Ruaa Ahmed, Ruaa Yasir, Safaa Awadallah, Sara Mohmmed, Suhaib Hassan [Ibrahim Malik Teaching Hospital, Khartoum]; Walid Shaban*, Aisha Hussein, Reem Rafea, Ahmed Abdalla, Abdalla Ahmed, Khalid Mohamed, Mansour Mohammed, Mohamed Altahir, Mohammed Adam, Omer Mohamed, Walaa Abdullah [Khartoum North Teaching Hospital (Bahri Hospital)]; Hammad Fadlalmola*, Ahmed Yassir Abdalla, Ahmed Ali Omer, Ahmed Alfatih Mustafa, Rawan Elhadi, Essam Eldien Abuobaida Banaga, Fatima Osman, Mohamed Abdalla, Hala Abdelhalim Mohamed Taha, Noon Ezzeldien Abdalmahmoud, Rofuida Hussien Nafie, Sami Jamal, Sharwany Ahmed [National Ribat University Hospital, Khartoum]

#### Syria

Rawan Alsheikh Ali*, Abdallah Aladna, Abdullah Aljoumaa, Hamdi Nawfal, Salma Jamali, Fatima Khouja, Ammar Niazi, Toka Al Rawashdeh [Aleppo University Hospital, Aleppo]

#### Tunisia

Nahla Kechiche*, Mouna Gara, Mouna Nasr, Marwen Baccar, Oumayma Benamor, Sawssen Chakroun [University Hospital Fattouma Bourguiba, University Of Monastir]

#### Turkey

Ahmet Necati Sanli*, Ahmet Yildiz, Mehmet Ali Demirkiran, Yildiz Buyukdereli Atadag, Yusuf Iskender Tandogan [Abdulkadir Yuksel State Hospital]; Esin Ozkan*, Yıldırım Ozer, Esin Ozkan, Muhammed Miran Oncel, Senad Kalkan [Bezmialem Vakif University, Faculty Of Medicine, Istanbul]; Tolga Gover*, Berke Manoglu, Ilayda Oksak, Ipek Kurt, Kerem Rifaioglu, Selman Sokmen, Tayfun Bisgin, Yasemin Yildirim, Abdil Yetkin Keskin [Dokuz Eylul Univ. Hospital, Izmir]; Tugce Dogan*, Berfin İlgaz Sahin, Cemil Aydin, Duygu Ece Benek, Hale Nur Tiras, Mert Arslangilay, Mert Aslangilay, Muhammet Yaytokgil, Mehmet Ali Capar, Yasemin Yazgan [Hitit University Faculty Of Medicine Çorum Research And Training Hospital]; Sebnem Bektas*, Ahmet Can Alagoz, Alara Ece Dagsali, Aylin Izgis, Kadir Uzel, Mustafa Soytas, Niyazi Cakir, Abdullah Emre Askin, Ibrahim Azboy, Kubilay Sabuncu, Merve Aslan, Melek Sahin, Mustafa Oncel, Nuri Okkabaz, Ramazan Kemal Sivrikaya, Alparslan Saylar, Dr. Alparslan Saylar, Meltem Yasar [Istanbul Medipol University Hospital, Istanbul]; Ergin Erginoz*, Haktan Ovul Bozkir, Kagan Zengin, Mehmet Faik Ozcelik, Server Sezgin Uludag, Zeynep Ozdemir [Istanbul University Cerrahpasa - Cerrahpasa School Of Medicine]; Osman Sibic*, Hatice Telci, Mehmet Abdussamet Bozkurt, Yasin Kara [Kanuni Sultan Suleyman Training And Research Hospital, Istanbul]; Mustafa Deniz Tepe*, Adnan Gündoğdu, Bilge Akın, Dilan Pehlivan, Ali Guner, Duygu Baysallar, Berkay Yıldız, Hale Cepe, Murat Emre Reis, Ayse Nilufer Yuzgec, Nurtac Kıralı, Taha Anıl Kodalak, Mehmet Ulusahin [Karadeniz Technical University Farabi Hospital, Trabzon]; Kamar Selim*, Ahmet Kale, Mehmet Emre Gecici, Melis Ozbilen [Kartal Dr. Lutfi Kirdar Training And Research Hospital, Istanbul]; Zeynep Düzyol*, Aylin Gemici, Elzem Korkmaz, Eminenur Şen, Muhammed Enes Taşcı, Elifsu Camkıran, Güşta Elieyioğlu, İkbal Kayabaş, Tevfik Kıvılcım Uprak, Canan Aral, Ayten Saraçoğlu, Mustafa Ümit Uğurlu, Zeynep Hazal Baltacı [Marmara University, School Of Medicine, Istanbul]; Ege Nur Akkaya*, Cem Fergar, Elif Zeynep Tabak, Guldane Zehra Kocyigit, Ilgaz Kayilioglu [Mugla Training And Research Hospital, Mugla]; Süleyman Polat*, Eli□f Çolak, Mehmet Emin Kara, Mert Candan, Mustafa Safa Uyanık, Ahmet Can Sarı [Samsun Training And Research Hospital, Samsun]; Attila Ulkucu*, Alperen Taha Certel, Arzu Dindar, Beyza Durdu, Cigdem Bayram, Eslem Kaya, Hakan Akdere, Ibrahim Ethem Cakcak, Ikranur Yavuz, Mert Omur, Mirac Ajredini, Erhan Onur Aydoğdu, Eylül Şenödeyici [Trakya University Faculty Of Medicine]; Ulku Ceren Koksoy*, Baturay Kansu Kazbek, Deniz Serim Korkmaz, Dogancan Yavuz, Hakan Yilmaz, Zeynep Sahan Cetınkaya, Elif Durmus, Filiz Tuzuner, Furkan Hokelekli, Mucahid Mutlu, Seyma Orcan Akbuz, Ziya Can Kus, Ziya Can Kus [Ufuk Üni□versi□tesi□ Tip Fakültesi□ Dr.Ri□dvan Ege Sağlik Araştirma Uygulama Merkezi□ Hastanesi□, Ankara]

#### United States of America

Michael Farrell*, Alayna Craig-Lucas, Matthew Painter, [Lehigh Valley Health Network]; Ashley Titan*, Aditya Narayan, Bunmi Fariyike, Lisa Knowlton, Tiffany Yue [Stanford Health Care, Palo Alto, California]; Emily Benham*, Abdelrahman Nimeri, Hope Werenski, Nicole Kaiser, Caroline Reinke [Atrium Health]

*Local lead

